# Bayesian Joint Model with Latent Time Shifts for Multivariate Longitudinal Data with Informative Dropout

**DOI:** 10.1101/2024.06.26.24309549

**Authors:** Xuzhi Wang, Martin G. Larson, Yorghos Tripodis, Michael P. LaValley, Chunyu Liu

## Abstract

Dementia often has an insidious onset with considerable individual differences in disease manifestation. Nonlinear mixed-effects models with latent time shifts have been proposed to investigate the long-term disease progression and individual disease stages. The latent time shift is a horizontal shift in time that aligns patients along a global timeline for disease progression. However, these models ignore informative dropout due to dementia or death, which may result in biased estimates of the longitudinal parameters. To account for informative dropout due to dementia or death, we propose a multivariate nonlinear joint model with latent time shifts. This joint model uses a multivariate nonlinear mixed-effects model with latent time shifts to model the correlated longitudinal markers of cognitive decline, and simultaneously, a proportional hazards model to incorporate dropout due to dementia or death. We investigate two association structures between the longitudinal process and the time to event process: the current value structure and the shared random effect structure. We compare the proposed joint model with separate models that ignore informative dropout across various simulation settings. The proposed joint models with correctly specified association structures show the best performance. Even the models with misspecified association structures outperform the separate models that does not consider informative dropout. We conclude that our proposed joint model with latent time shifts offers more accurate and robust estimates than the latent time disease progression models that neglect informative dropout. Future research will involve incorporating competing risks and other parametrizations of the longitudinal model into this joint model framework.

## 1 Introduction

Dementia is characterized by a decline in mental ability that impairs memory, thinking, and decision-making abilities, which significantly affects people’s daily activities [1]. It is reported that dementia serves as the leading cause of dependence and disability among the elderly worldwide [2]. Modeling disease progression using cognitive decline preceding dementia onset is challenging since dementia often has an insidious onset with considerable individual differences in disease manifestation [3]. Methods have been proposed to describe the progression of dementia including linear or non-linear mixed-effects models [4–7]. However, traditional timescales used in these models such as time to diagnosis, time since inclusion, or chronological age face challenges in accurately depicting the progression of dementia [8]. Specifically, the use of time to diagnosis can be unreliable and subjective, and it confines the analysis to only those participants who have developed dementia, consequently leading to a reduction in sample size. Time since inclusion lacks biological meaning and exhibits heterogeneity due to varying clinical stages at which participants are enrolled in the study. Chronological age introduces considerable individual heterogeneity as disease manifests at different ages for different people. Besides, the substantial individual differences in disease manifestation pose challenges in accurately staging patients.

Due to the limitations of traditional timescales, the true longitudinal trajectory for disease progression may be obscured using these timescales. Therefore, disease progression models that utilize a latent disease time have been proposed [3, 9–11]. The latent disease time is retrieved from the data using time-warping functions by estimating a subject-level latent time shift for individual heterogeneity in disease stages. The subject-level latent time shift realigns participants along a global timeline in the population by assuming that participants experience overall the same disease progression. Compared with the mix-effects models based on traditional timescales, the disease progression models offer the following benefits. First, the latent disease time serves as a continuous representation of the disease stage by re-ordering the participants according to disease severity, which may provide a more accurate staging of participants compared to coarse disease-stage groups [3]. Second, these models aid in characterizing the temporal order of the occurrence of different cognitive changes since all cognitive measures can be compared on the global timescale. Third, these models can effectively recover the true long-term disease progression trajectory from short-term observations [9]. However, the disease progression models differ in how they parameterize latent time shifts, the shapes of mean disease progression trajectories, and the number of cognitive measures they can model simultaneously. Donohue et al. (2014) proposed a semiparametric model and iterative estimation procedure to estimate the long-term disease trends based on short-term data [10]. Li et al. (2019) developed a latent time joint mixed model using Bayesian estimation [9]. Multiple longitudinal outcomes were simultaneously modeled, and the nonlinear disease progression trends were accommodated by a nonlinear transformation prior to the trajectory modeling. Based on Li et al. (2017), Lespinasse et al. (2023) further used the partially observed clinical diagnosis information to help anchor the estimation of latent time shifts [8]. Raket et al. (2020) directly modeled a nonlinear longitudinal cognitive trajectory on its original scale by a parametrized family of exponential functions and included covariate effects on disease stage, rate of decline and deviation from the mean [11]. Based on Raket et al. (2020), Kühnel et al. (2021) simultaneously modeled multivariate longitudinal trajectories that may differ in sensitivity across disease stages by proposing a multivariate continuous-time disease progression (MCDP) model [3].

One common limitation in these existing disease progression models is their inability to accommodate informative dropout. In longitudinal studies tracking dementia progression, dropout is often caused by the occurrence of dementia or death [12]. When the probability of a subject dropping out depends on their unobserved longitudinal measurements, e.g., cognitive decline, this type of dropout process is identified as nonrandom or informative, or missing not at random (MNAR) [13]. However, the latent disease progression models usually assume that the missing data are missing at random (MAR), which indicates that the missingness can be fully accounted for using the observed data. In order to obtain valid inference, a model for the joint distribution of the longitudinal and missingness processes should be considered. Selection models, pattern-mixture models, and joint models, also known as shared parameter models, have been proposed to handle informative dropouts [14–17]. Cuer et al. (2020) compared these three strategies for informative missingness [18]. Compared with selection models, pattern-mixture models and joint models allow researchers to test the mechanism of missing data. In addition, joint models treat time-to-dropout as continuous and considers dropouts related to different clinical events. Joint models provide a framework to jointly model both longitudinal and missingness processes by postulating a mixed effect model for the longitudinal outcome and a survival model for the time-to-event outcome (e.g., time to dropout due to dementia or death) [19]. Both processes in the joint model framework are linked by a set of random effects that are assumed to capture the associations between these two outcomes. A joint model is also essential in scenarios where the objective is to assess the association between longitudinal and time-to-event processes [19]. Various types of joint models have been proposed in terms of estimation approaches (e.g., frequentist or Bayesian [20]), the number of longitudinal outcomes (e.g., univariate or multivariate [21]), the association structures between longitudinal and survival processes (e.g., current value or shared random effects [19]), the parametrization of the longitudinal sub-model (e.g., linear or nonlinear [22]), and the types of time-to-event outcomes (e.g., single event, multiple events, or recurrent events [23]).

To demonstrate the capability of joint models to handle informative missingness, researchers have compared joint models with mixed-effects models that treat informative missingness as MAR, and with separate models of longitudinal and missingness processes without accounting for the association between them. Among these studies, joint models employing the shared random effect association structure have been used, where dropouts are assumed to be directly related to the random effects from the longitudinal process [24–27].

Another commonly used association structure is the current value structure, where dropouts are linked to the true unobserved values of the longitudinal trajectories [18, 28–32]. Notably, Vonesh et al. (2006) investigated both association structures, comparing them with separate models under various assumptions of missing mechanism [17]. All studies showed that joint models improve over linear mixed models or separate models regarding bias and model fit. In addition, it has been shown that linear mixed models tend to provide overly optimistic estimates for longitudinal outcomes, while joint models are able to recover the true longitudinal estimates with low bias [26–28]. When dropouts occur due to different reasons, joint models with competing risks have been developed to account for various types of dropouts. Elashoff et al. (2008) and Williamson et al. (2008) assessed joint models with competing risks based on the shared random effect structure against separate models that ignore dropouts [25, 33]. Gueorguieva et al. (2012) further extended the previous two studies to allow for interval-censored dropout times and incorporated a joint model treating all dropouts equally into the comparative analysis [26].

Results indicated that the joint model with competing risks due to different reasons outperformed the joint model with a common dropout reason. The difference between two joint models was more pronounced in the scenario where one type of dropouts was informative, and the other one was non-informative.

In this project, we aim to describe a multivariate nonlinear joint model with latent time shifts. Specifically, the joint model combines a model to consider a nonlinear latent time disease progression with multiple longitudinal measures and a proportional hazards model to consider informative dropout using Bayesian methods. To demonstrate the ability of our proposed model to account for informative dropout, we also aim to compare the proposed joint models based on different association structures with the disease progression model that ignores informative dropout. Furthermore, we apply our proposed joint model to the Framingham Heart Study (FHS) Offspring Cohort data.

## 2 Methods

Our proposed joint model is an extension of the multivariate continuous-time disease progression (MCDP) model proposed by Kühnel et al. (2020) [3]. The MCDP model is a multivariate nonlinear mixed-effects model that aligns patients according to their predicted disease progression along a global latent timescale. Compared to other disease progression models that utilize latent disease time, the MCDP model demonstrates superior performance in terms of more accurate predictive postbaseline trajectories and individual patient staging based on predicted subject-level latent time shifts. Therefore, the estimated disease progression trajectories in the MCDP model more accurately reflect the true evolution of cognitive measures preceding dementia and enhance predictions of individual disease progression. Overall, our proposed joint model is composed of three components: (i) the MCDP model for longitudinal cognitive measures (longitudinal sub-model); (ii) a proportional hazards model for the missingness process, i.e., the time to dementia or death (survival sub-model); (iii) an association structure that links processes (i) and (ii).

### 2.1 Longitudinal sub-model: MCDP

Let *Y*_*k*i_(*t*_ij_) be the value of the *k*th longitudinal cognitive measure at time *t*_ij_ for subject *i*, where *i* = 1, 2, …, *N*, *k* = 1, 2, …, *K*, and *j* = 1,2, …, *n*_*k*i_. The following parameters in bold indicate vectors. For participant *i*, let *Z*_*i*_ be a vector of length *p* for baseline covariates and *X*_*i*_ be a vector of length *q* for covariates that have an influence on disease stages. The MCDP model can be formulated as

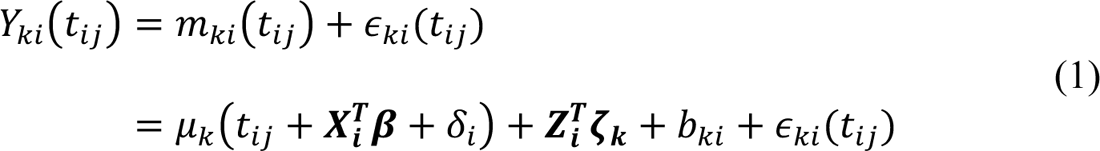

where *m*_*k*i_5*t*_ij_6 denotes the true unobserved value of *Y*_*k*i_5*t*_ij_6 without the random error ∈_*k*i_(*t*_ij_), with ∈_*k*i_(*t*_ij_)∼*N*(0, σ^2^). μ_*k*_ is a function for the mean disease progression in the population. An individual’s deviation from the mean disease progression can be divided into two parts: the horizontal shift *X*_*i*_^*T*^β + δ_i_ and the vertical shift *Z*_*i*_^*T*^ॐ_*k*_ + *b*_*k*i_ + ∈_*k*i_(*t*_ij_).

#### Horizontal shift

The horizontal shift refers to the latent time shift that moves subjects backward or forward in time according to their true disease stages, so subjects are realigned along a global timeline. The latent time shift is composed of a fixed effect *X*^*T*^β for individual differences in disease stage that can be ascribed to covariates *X*_*i*_, and a random effect δ_i_ that represents the unobserved random variation in disease stage with δ_i_∼*N*(0, σ^2^). The timescale formed by the shifted time points *t*_ij_ + *X*^*T*^β + δ_i_, also referred to as the ‘disease time’ or ‘disease age’, constitutes a global timescale for disease progression in the population. If the original timescale *t*_ij_ is chronological age and (*X*^*T*^β + δ_i_) < 0, a participant’s disease age is younger than his/her chronological age (**Figure 1**). That is, the participant’s disease stage is earlier than the average disease stage for his/her actual chronological age. The subject-specific latent time shift is assumed to be consistent across all *K* outcomes for each participant. Previous studies revealed that the common individual disease stage assumption is feasible in the presence of multiple outcomes that may differ in sensitivity across disease stages [3]. The horizontal latent time shift is visualized in **Figure 1**.

**Figure 1.**
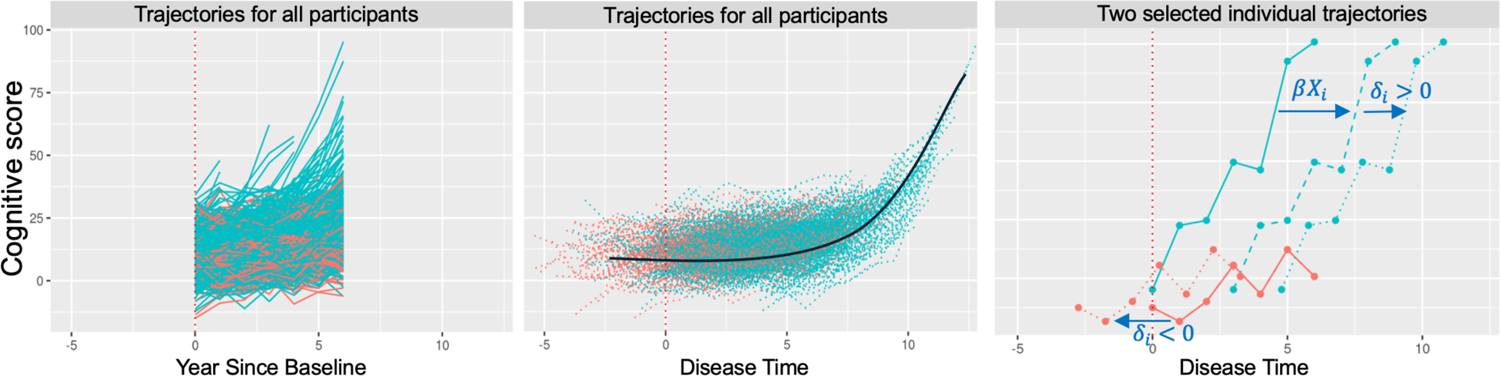
Simulated trajectories with years since baseline and adjusted disease time. The three panels illustrate cognitive score trajectories, all using cognitive score as the y-axis. The left panel plots trajectories against the original years since baseline (*t*_ij_), showing all participant data. The middle panel adjusts the x-axis to disease time (*t*_ij_ + β*X*_i_ + δ_i_) and realigned all trajectories in the first panel based on this adjusted timeline. Blue and red lines/dots depict participants with *X*_i_ = 1 and *X*_i_ = 0, respectively. The solid black curve in this panel marks the predicted mean trajectory using the adjusted disease time. The right panel shows trajectories for two specific individuals against disease time. For one individual, his/her blue solid trajectory shifts to a dashed trajectory after adjusting for a fixed time shift (β*X*_i_), then to a dotted trajectory with a further random time shift (δ_i_), *i* = 1. Similarly, the red solid trajectory for another individual shifts left to a dotted line after accounting for a random time shift (δ_i_) only, *i* = 2. Vertical shifts are the deviations of each individual trajectories from the predicted mean trajectory, which is not shown in the figures.

#### Vertical shift

The vertical shift represents a deviation from the mean disease progression and is composed of three parts: a fixed effect *Z*^*T*^ॐ_*k*_, which accounts for the deviation that can be attributed to covariates *Z*_*i*_; a random intercept *b*_*k*i_, which captures the subject-level unobserved random deviation; and a random error term ∈_*k*i_(*t*_ij_). The random intercept *b*_*k*i_ resembles the random intercept in the traditional linear mixed-effects model. We assume the vector of random effects across *K* longitudinal measures, denoted as *b*_*i*_ = (*b*_1i_, *b*_2i_, …, *b*_*K*i_)′, follows a multivariate normal distribution: *b*_*i*_∼*MVN*(0, Σ_*b*_). (σ^2^_b1_, σ^2^_b2_, …, σ^2^_bk_)^*T*^ is the variance vector on the diagonal of Σ_*b*_.

We further assume independence among the random latent time shift δ_i_, the random intercept *b*_*i*_, and the random error term ∈_*k*i_(*t*_ij_).

#### Mean disease progression

To model the mean disease progression trajectory for the *k*th longitudinal outcome, we use a nonlinear form for μ_*k*_:

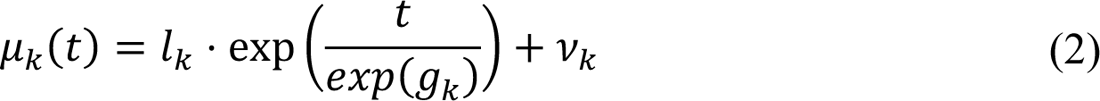

where *v*_*k*_ is an intercept parameter describing the left asymptote; *l*_*k*_ is a scaling parameter for the exponential function, which can be interpreted as the mean deviation from *v*_*k*_ at time *t* = 0. *g*_*k*_ is a scaling parameter for time. The decision to use an exponential mean curve is based on the observation that cognitive decline in dementia is typically monotonic over the long run. Besides, cognitive decline is generally characterized by an escalating rate of deterioration over the course of disease progression, which is consistent with the shape of an exponential curve [3].

### 2.2 Survival sub-model: proportional hazards model

During follow-up, a subject may drop out of the study due to dementia or death. We use a parametric proportional hazards model for the time to dropout due to dementia or death. The model is as follows

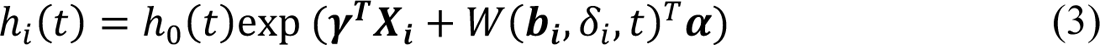

where ℎ_-_(*t*) is the baseline hazard. It has been shown that an unspecified baseline hazard in the joint model is likely to result in the underestimation of the standard errors of the parameter estimates [34]. To avoid this issue, we use a parametric Weibull baseline hazard. It can be extended to other parametric forms such as exponential or piecewise constant. γ is the vector of coefficients for covariates *X*_*i*_. *W*(*b*_*i*_, δ_i_, *t*) is a function of random effects from the longitudinal sub-model (i.e., random intercepts *b*_*i*_ and random time shift δ_i_) and time *t*, which represents the association structure between the longitudinal and survival (e.g., missingness or dropout) processes. α denotes the associated coefficients for *W*(*b*_*i*_, δ_i_, *t*). Specifically, we consider two commonly used forms for *W*(*b*_*i*_, δ_i_, *t*): the current value structure and the shared random effect structure. In addition, to facilitate model comparison we include the separate models that ignores the association between the longitudinal and survival processes, where α=0.

#### Current value association structure (CV)

This association structure assumes that the true unobserved value for longitudinal measures at time *t*, denoted as *m*_*k*i_(*t*), is predictive of the event risk at the same time *t* [21]. Therefore, we have *W*(*b*, δ, *t*) = 5*m* (*t*), *m* (*t*), …, *m* (*t*)6^*T*^. Equation (3) can be rewritten as

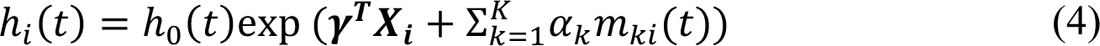

where α_*k*_ measures the association between *m*_*k*i_(*t*) and the risk for the event of our interest (e.g., dropout due to dementia or death), indicating that each one-unit increase in the current value of *m*_*k*i_(*t*) at time *t* is associated with an exp (α_*k*_)-fold increase in the hazard of the event at the same time *t*. This particular association structure is often employed in scenarios where the primary focus is on survival time. After removing the error term, the longitudinal sub-model is considered a time-dependent covariate in the survival process [34].

#### Shared random effect association structure (RE)

In real-world scenarios, it’s not always realistic to assume a current value association structure. The shared random effect association structure (RE) postulates that the risk for the event of our interest is directly associated with the random effects in both horizontal and vertical shifts from the longitudinal sub-model, i.e., the random time shift δ_i_ and the random intercepts *b*_*i*_. We have *W*(*b*_*i*_, δ_i_, *t*) = (δ_i_, *b*_1i_, *b*_2i_, …, *b*_*K*i_)^*T*^. Equation (3) can be rewritten as

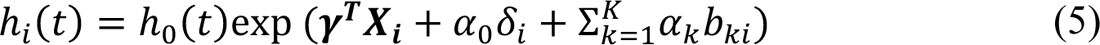

where α_-_ and α_*k*_ measure the strength of the association between the event risk and the random latent time shift as well as the random intercepts, respectively. Consider two individuals with similar demographics but different disease stages as an example. If we assume α_-_ > 0 and α_*k*_ > 0, the individual with an earlier disease stage (i.e., a smaller random latent time shift δ_i_) or exhibit a smaller baseline value (i.e., smaller *b*_*k*i_) is at a lower risk of the event at a given time *t* after adjusting for other factors. Typically, this structure is utilized in scenarios where the primary focus is the longitudinal process with informative dropout, or in situations where both the longitudinal and survival processes are of equal interest [34].

#### Separate models

Note that if we set α_*k*_ = 0, *k* = 1,2, …, *K*, in the current value association structure, or α_-_ = α_*k*_ = 0, *k* = 1,2, …, *K*, in the shared random effects association structure, the survival sub-model is reduced to a standard proportional hazards model, i.e.,

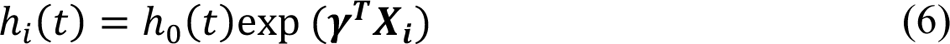

Since this model fails to accommodate the association between the longitudinal and survival processes, the joint model is reduced to the separate models of the longitudinal and survival processes. That is, the longitudinal and survival processes are modeled independently in the separate models. The estimates from the longitudinal MCDP model in the separate models are equivalent to those obtained from a standard MCDP model that ignores informative dropout.

### 2.3 The joint model overview

We use **Figure 2** to illustrate our proposed nonlinear joint model framework assuming there are two longitudinal cognitive measures (*K* = 2). The random effects including the random latent time shift, denoted as δ_i_, and the random intercepts, denoted as *b*_1i_ and *b*_2i_. These parameters play important roles in constructing the joint model. First, the random effects capture the correlation between the longitudinal measures. Second, the longitudinal and survival processes are linked directly through the random effects in the shared random effect model. In contrast, the random effects have an indirect effect on the hazard via the true unobserved values of the longitudinal measures, *m*_1i_(*t*) and *m*_2i_(*t*), in the current value model

**Figure 2.**
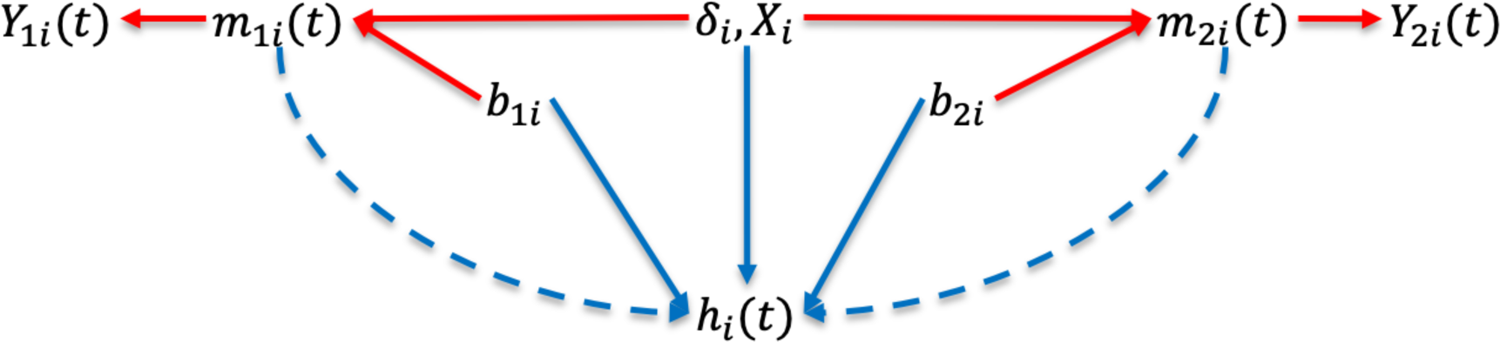
The joint model framework. *Y*_1i_ and *Y*_2i_ are two longitudinal measures. *m*_1i_(*t*) and *m*_2i_(*t*) are the true unobserved values for two longitudinal measures. δ_i_ is the random latent time shift and *X*_i_ is a vector of covariates. *b*_1i_ and *b*_2i_ are correlated random intercepts for the first and second longitudinal measures. ℎ_i_(*t*) denotes the hazard function for the survival process. Both longitudinal and survival processes are linked by either random effects (solid blue lines for the shared random effect model) or the true unobserved values of longitudinal measures (dashed blue lines for the current value model).

### 2.4 Estimation of the joint model

#### 2.4.1 Likelihood

Let *T*^∗^ be the true time to dropout due to dementia or death for subject *i*, and *C*_i_ the censoring time, which corresponds to dropout due to other reasons. Let *T*_i_ = min {*T*^∗^, *C*_i_} denote the observed time. Δ_i_ = min {*T*^∗^, *C*_i_} is the indicator for death. Let ψ denote the full parameter vector. δ_i_ and *b*_*i*_ are the random latent time shift and the vector of random intercepts. The likelihood is constructed under the assumption that both the longitudinal and survival processes are independent given the random effects. Therefore, the likelihood conditional on the random effects is as follows:

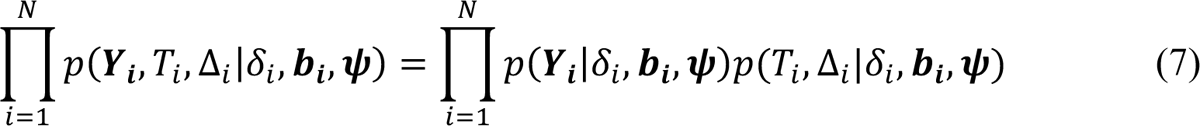

For the longitudinal part, we have

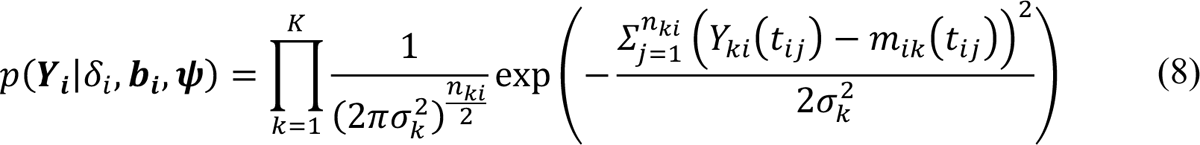

For the survival part, we have

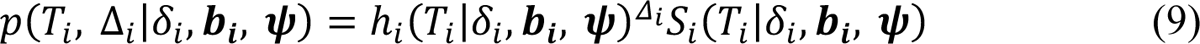

where *h*_i_(*t*) is the hazard function described in Equations (4) to (6). *S*_i_(*T*_i_|δ_i_, *b*_*i*_, ψ) = exp (−∫ ^*T*i^*h* (*s*|δ, *b*_i_, ψ)*ds*), which represents the survival function. Note that we use Gauss-Kronrod quadrature [35] with 15 nodes for numerical approximation of integration when the survival sub-model is a current value model. Specifically, 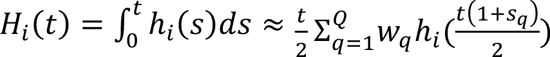, in which *w_q_* is the standardized weight for quadrature node *q* with *q* ∈ {1,2, …, *Q*}. We set *Q* = 15. *s*_*q*_ is the location for quadrature node *q*.

#### 2.4.2 Bayesian Inference

We use a Bayesian approach based on Markov Chain Monte Carlo (MCMC) methods for the parameter estimation of the joint model. For the Bayesian approach, the random effects, including the random latent time shift and random intercepts, are treated as model parameters. The joint posterior probability distribution is analogous to:

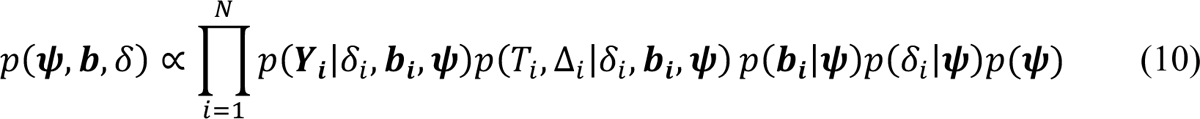

where *p*(ψ) is the joint prior distribution for the full parameter vector ψ. *p*(δ_i_|ψ) and *p*(*b*_*i*_|ψ) are the distributions for the random latent time shift and random intercepts with normal distributions, respectively.

We use weakly informative priors *p*(ψ) on all the model parameters to allow the data to determine the estimation of parameters. For the fixed effects in the longitudinal sub-model, i.e., *g*_*k*_, *v*_*k*_, β, ॐ_*k*_, we assign normal priors with a large variance, i.e., *N*(0, 100). Note that for the scaling factor *l*_*k*_, we use a uniform prior with an upper limit of 0, denoted as *Uniform*(−1, 0), to ensure that all cognitive trajectories are oriented to be decreasing (*l*_*k*_ < 0), consistent with real-world scenarios. The fixed parameters in the survival sub-models, denoted as γ and α, are also assigned weakly informative normal priors *N*(0, 100). We use a Half-Cauchy (0, 2.5) prior for the standard deviation of the random effects, including σ_δ_ for the latent time shift, σ_*bk*_ for the random intercepts, and σ_*k*_ for the random errors across longitudinal measures, *k* = 1,2, …, *K*.

The variance-covariance matrix for the random intercepts (Σ_*b*_) is decomposed into a correlation matrix (Ω_*b*_) and two diagonal matrices (Λ_*b*_) with standard deviation terms on the diagonal using Cholesky decomposition, i.e., Σ_*b*_ = Λ_*b*_Ω_*b*_Λ_*b*_. We employ the “Lewandowski-Kurowicka-Joe (LKJ) priors” [36] with a shape parameter equal to 2, i.e., the parameterization of the LKJ correlation matrix density in terms of its Cholesky factor, for estimating the correlation matrix Ω_b_.

The model parameters are estimated using the R interface of Stan with the R package “rstan” [37]. Stan utilizes a No-U-Turn Sampler version of Hamiltonian Monte Carlo (HMC) algorithm which optimizes the exploration of the target distribution based on Hamiltonian dynamics [38]. It has been shown that HMC has the capacity to handle complex nonlinear estimation tasks that might prove challenging for Gibbs sampling [39]. To assess model convergence, we use the potential scale reduction statistic Ř calculated by Stan [40]. Ř values < 1.1 for all parameters indicate successful model convergence.

Model comparison is evaluated using the Widely Applicable Information Criterion (WAIC) [41], also known as the Watanable-Akaike for Bayesian model selection. WAIC is a useful tool for estimating pointwise out-of-sample prediction accuracy within the Bayesian framework. It is calculated based on the log-likelihood evaluated at the posterior simulations of the parameter values. Compared to the deviance information criterion (DIC), which is another commonly used measure in Bayesian methods, WAIC is generally more stable and is considered an improvement over DIC [42]. A smaller value of WAIC indicates a better model fit.

## 3. Simulation study design

Under various scenarios, we evaluated and compared the joint models based on the shared random effect and current value structures. We also compared the joint models with the separate models.

### 3.1 Data generation

We generated a cohort with n=1000 subjects. We also generated a baseline time variable *t*_i-_ from a uniform distribution *U*(−5, 5) to be the centered age around 60 years (i.e., the real age subtracted by 60) at the baseline. Each participant was examined every four years until the 20^th^ year in the follow-up. The longitudinal measures were observed until the occurrence of an event or at the 20^th^ year of the follow-up period. We employed the MCDP model to generate these two longitudinal measures, *Y*_1i_5*t*_ij_6 and *Y*_2i_5*t*_ij_6, based on the following two equations:

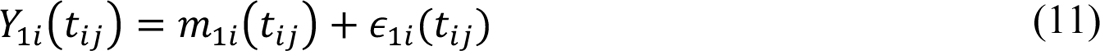

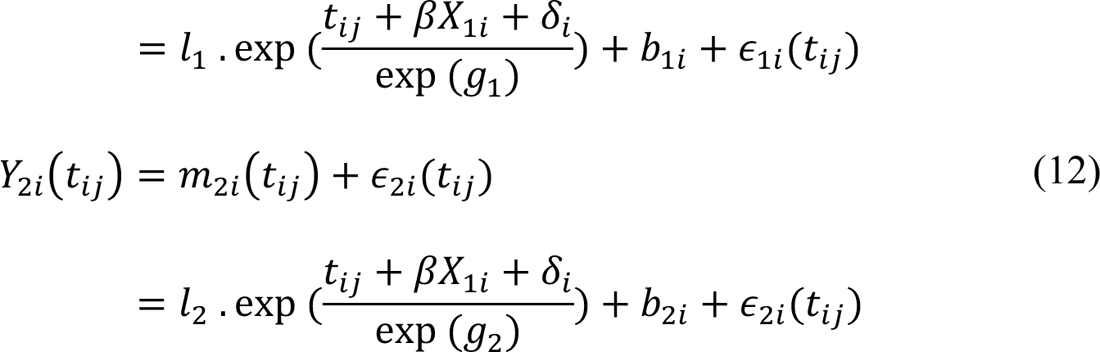

We considered one binary covariate *X*_1i_ that was generated from Bernoulli (0.5). *X*_1i_ had an impact on the latent time shift and no covariates were included in the vertical shift since our focus was on examining the trajectories of the fitted longitudinal measures over time. The correlation between two longitudinal measures were captured by both the random latent time shift and the random intercepts. We assume that the random latent time shift followed a normal distribution, i.e., δ_i_∼*N*(0, σ^2^), and the random intercepts had the following distribution

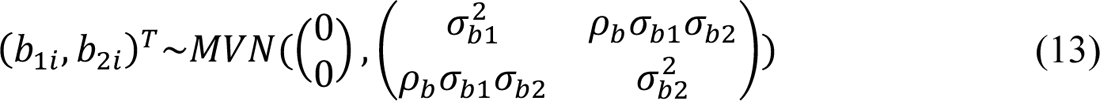

We simulated the dropout times due to dementia or death under three association structures described in the Methods section: the shared random effects structure, the current value structure, and the separate models. When the current value structure was implemented, the event times were generated based on the following equation

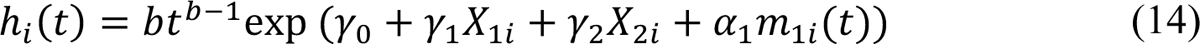

In addition to the binary covariate *X*_1i_, which was shared with the longitudinal sub-model, we also included a continuous covariate *X*_2i_ generated from *Uniform* (−5, 5). We incorporated the true unobserved value of the first longitudinal measure at time *t* to capture the association between two processes. In the survival sub-model, the true unobserved value of the second longitudinal measure was excluded to avoid multicollinearity assuming two longitudinal measures were moderately or highly correlated. After specifying the proportional hazards model, the event times were then generated using an inverse cumulative hazard function transformation [43]. Specifically, we first simulated a variable *u*_i_∼*Uniform* (0,1) for participant *i*. Next, we obtained the true event time *T*^∗^ by solving for the equation *u*_i_ = *S*_i_(*t*), where *S*_i_(*t*) is survival function in Equation (9). We then defined the censoring time, *C*_i,_ as the time to the maximum follow-up duration for participant *i*. Lastly, the observed event time was *T*_i_ = min {*T*^∗^, *C*_i_}. For participant *i*, the longitudinal measures, *Y*_1i_5*t*_ij_6 and *Y*_2i_5*t*_ij_6, occurred after *T*_i_ were dropped.

In the context where the shared random effect structure was our primary interest, the event times were generated based on the following equation

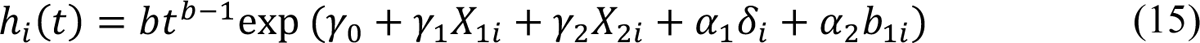

We incorporated the random latent time shift δ_i_ and the random intercept *b*_1i_ from the first longitudinal measure to capture the association between two processes. Like the current value data structure, we mitigated multicollinearity by excluding the random intercept *b*_2i_ from the second longitudinal measure in the survival sub-model. The event times can be generated following the procedures outlined in the current value structure or directly from a Weibull distribution based on the hazard function as specified in Equation (15).

In contrast with the previous two association structures, we also considered a scenario where the dropout times were not related to the longitudinal measures, implying non-informative missingness. The non-informative dropout times were generated from the following proportional hazards model with the independent association structure between the longitudinal and survival processes (survival sub-model in the separate models)

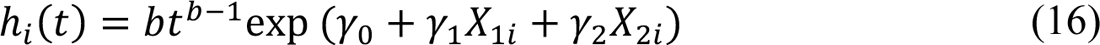

where only *X*_1i_ and *X*_2i_ were included as covariates. The event times generation procedure was the same as the shared random effect structure.

### 3.2 Simulation scenarios

Our simulated dataset mimicked the Offspring cohort in the FHS to allow for reasonable generalization. We generated 6 scenarios and summarized all parameter settings in **Table 1**. Three association structures described in the previous section were considered to reflect various potential real-world conditions about the relationships between the longitudinal and survival processes. Scenario 1 (the shared random effect structure) indicated that a participant at a later disease stage and smaller baseline cognitive values was more likely to dropout. Scenario 2 (the current value structure) implied that a higher hazard for dropout was associated with lower unobserved current values of longitudinal cognitive measures. In Scenario 3 (separate structure), the dropout times were only dependent on the observed covariates *X*_1i_ and *X*_2i_, unrelated to the values of longitudinal measures. Scenarios 4, 5, and 6 were based on the shared random effect association structure, resembling Scenario 1. In Scenario 4, we evaluated the impact of a smaller sample size on model performance by setting n=500. In Scenario 5, the event rate was increased from 30% to 50% by varying the baseline hazard parameters. In Scenario 6, the latent time shift was assumed to have a longer span. For each scenario, 200 datasets, each with 1000 subjects (except Scenario 5 with n=500), were generated and analyzed.

**Table 1.** Parameter settings for all 6 scenarios in the simulation study.

| Parameters | Scenario 1 | Scenario 2 | Scenario 3 | Scenario 4 | Scenario 5 | Scenario 6 |
| --- | --- | --- | --- | --- | --- | --- |
| **Longitudinal** |  |  |  |  |  |  |
| N sample size | 1000 | 1000 | 1000 | 500 | 1000 | 1000 |
| $l_1$ | -0.06 | -0.06 | -0.06 | -0.06 | -0.06 | -0.06 |
| $g_1$ | 2.15 | 2.15 | 2.15 | 2.15 | 2.15 | 2.15 |
| $v_1$ | 0.3 | 0.3 | 0.3 | 0.3 | 0.3 | 0.3 |
| $l_2$ | -0.02 | -0.02 | -0.02 | -0.02 | -0.02 | -0.02 |
| $g_2$ | 2.1 | 2.1 | 2.1 | 2.1 | 2.1 | 2.1 |
| $v_2$ | 0.15 | 0.15 | 0.15 | 0.15 | 0.15 | 0.15 |
| $\beta$ | -2 | -2 | -2 | -2 | -2 | -2 |
| $\sigma_\delta$ | 8.5 | 8.5 | 8.5 | 8.5 | 8.5 | 4 |
| $\sigma_{b1}$ | 0.45 | 0.45 | 0.45 | 0.45 | 0.45 | 0.45 |
| $\sigma_{b2}$ | 0.5 | 0.5 | 0.5 | 0.5 | 0.5 | 0.5 |
| $\rho_b$ | 0.6 | 0.6 | 0.6 | 0.6 | 0.6 | 0.6 |
| $\sigma_1$ | 0.3 | 0.3 | 0.3 | 0.3 | 0.3 | 0.3 |
| $\sigma_2$ | 0.2 | 0.2 | 0.2 | 0.2 | 0.2 | 0.2 |
| **Survival** |  |  |  |  |  |  |
| $b$ | 4.9 | 2.9 | 3.7 | 4.9 | 5 | 4.9 |
| $\gamma_0$ | -16 | -10 | -12 | -16 | -15 | -16 |
| $\gamma_1$ | -0.5 | -0.2 | -0.5 | -0.5 | -0.5 | -0.5 |
| $\gamma_2$ | -0.05 | -0.05 | -0.1 | -0.05 | -0.05 | -0.05 |
| $\alpha_1$ | 0.2 | -1 | NA | 0.2 | 0.2 | 0.2 |
| $\alpha_2$ | -0.1 | NA | NA | -0.1 | -0.1 | -0.1 |

Scenario 1, shared random effect association structure; Scenario 2, current value association structure; Scenario 3, separate (independent) association structure; Scenario 4, shared random effect association structure with N=500; Scenario 5, shared random effect association structure with a higher event rate of 50%; Scenario 6, shared random effect association structure with a smaller span of latent time shift of 4.

### 3.3 Analysis models

For each scenario, we used the following three models to analyze the data. We used the MCDP model described in Equations (11) and (12) as the longitudinal sub-model, while we consider three different survival sub-models: (i) Model 1 (MCDP+RE): the shared random effect association structure in Equation (14); (ii) Model 2 (MCDP+CV): the current value association structure in Equation (15); (iii) Model 3 (separate models): the separate analysis of longitudinal and survival processes in Equation (16); In Bayesian estimation, we used weakly informative priors described in the Methods section and obtained 6000 MCMC sample iterations from each of the 3 parallel chains with the first 4000 iterations as a warm-up phase. Convergence was evaluated through the potential scale reduction statistic Ř.

## 4. Simulation results

The objectives of our simulation study were threefold. First, to show our proposed joint models are capable of realigning participants along a global disease time. Second, to demonstrate our proposed joint models effectively account for informative dropout by retrieving the true mean disease progression trajectory. Third, to illustrate that the estimated latent time shifts from our proposed joint models were more accurate than those derived from the MCDP model in the separate models, which ignores informative dropout. In the simulation results, we focused on the following parameters of the interest: the scaling parameters for the exponential curve *l*_)_, *g*_)_, *l*_2_, and *g*_2_, the time shift parameters β and σ_δ_, as well as the association parameters α_=_ and α_>_ estimated in the shared random effect and current value models.

### Scenario 1. Shared random effect data structure (Table 2)

Figure 3 showed an example of one simulated dataset of 1000 subjects who were followed up to 20 years with up to six longitudinal measures. The two panels in the “Observed” column represented the simulated individual trajectories for two longitudinal measures on the original timescale without considering latent time shifts. No clear trajectory patterns were observed since the true mean trajectories were obscured on the original timescale. After applying the MCDP model in the separate models and our proposed joint models (panels in the “Separate”, “MCDP+RE”, and “MCDP+CV” columns in Figure 3), the participants were realigned according to a global disease timeline, allowing the true disease progression trajectories to be reflected in the realigned data.

**Figure 3.**
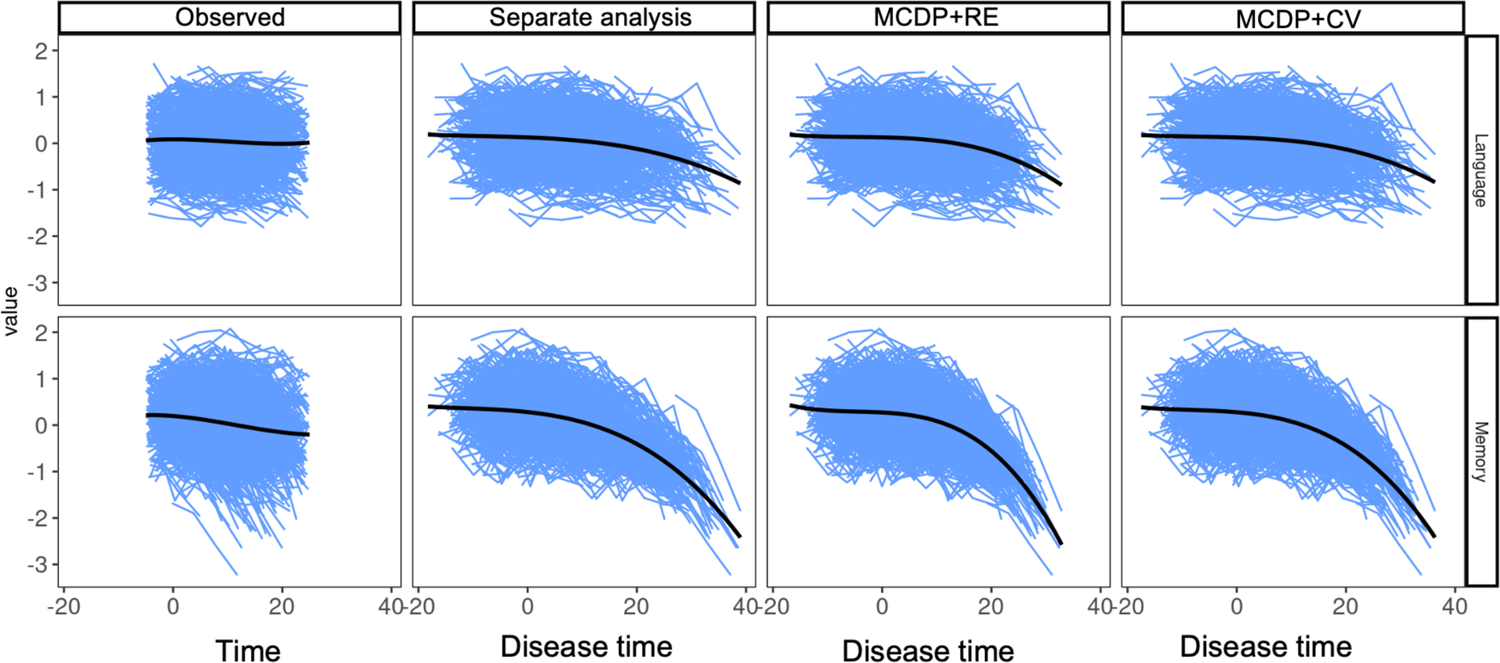
One simulated example of trajectories on the original time and disease time. The eight panels illustrate the simulated trajectories for two cognitive measures. The y-axis in each panel denotes the value of cognitive measures, and the x-axis represents the simulated time. The two panels in the first column denotes the trajectories aligned on the original timescale. The other panels represent the trajectories that are realigned by fitting different models with latent time shifts. MCDP+RE, the joint model with the shared random effect association structure; MCDP+CV, the joint model with the current value association structure.

**Table 2.** Simulation results for Scenario 1 (shared random effect association structure).

| Parameter | True | Separate analysis<br>(WAIC=4993) |  |  | MCDP+RE<br>(WAIC=4468) |  |  | MCDP+CV<br>(WAIC=4726) |  |  |
| --- | --- | --- | --- | --- | --- | --- | --- | --- | --- | --- |
|  |  | Means | Bias | CP | Means | Bias | CP | Means | Bias | CP |
| Longitudinal |  |  |  |  |  |  |  |  |  |  |
| $l_1$ | -0.06 | -0.09 | 48.36 | 0.21 | -0.06 | 5.86 | 0.95 | -0.07 | 18.88 | 0.82 |
| $g_1$ | 2.15 | 2.39 | 11.22 | 0.01 | 2.17 | 0.70 | 0.92 | 2.27 | 5.63 | 0.39 |
| $v_1$ | 0.30 | 0.33 | 9.12 | 0.76 | 0.30 | 1.45 | 0.98 | 0.31 | 3.16 | 0.94 |
| $l_2$ | -0.02 | -0.03 | 55.45 | 0.46 | -0.02 | 8.21 | 0.92 | -0.03 | 29.11 | 0.81 |
| $g_2$ | 2.10 | 2.34 | 11.42 | 0.07 | 2.12 | 0.78 | 0.93 | 2.24 | 6.82 | 0.40 |
| $v_2$ | 0.15 | 0.16 | 8.42 | 0.91 | 0.15 | 2.21 | 0.94 | 0.16 | 4.50 | 0.93 |
| $\sigma_1$ | 0.30 | 0.30 | 1.05 | 0.85 | 0.30 | 0.22 | 0.95 | 0.30 | 0.27 | 0.93 |
| $\sigma_2$ | 0.20 | 0.20 | 0.23 | 0.97 | 0.20 | 0.00 | 0.98 | 0.20 | 0.09 | 0.98 |
| $\sigma_{b1}$ | 0.45 | 0.45 | 0.01 | 0.94 | 0.45 | -0.29 | 0.93 | 0.45 | -0.06 | 0.93 |
| $\sigma_{b1}$ | 0.50 | 0.50 | -0.31 | 0.95 | 0.50 | -0.40 | 0.95 | 0.50 | -0.22 | 0.94 |
| $\rho_b$ | 0.60 | 0.60 | 0.08 | 0.94 | 0.60 | -0.20 | 0.93 | 0.60 | 0.00 | 0.94 |
| Time shift |  |  |  |  |  |  |  |  |  |  |
| $\beta$ | -2.00 | -2.20 | 9.82 | 0.93 | -2.07 | 3.73 | 0.96 | -2.15 | 7.26 | 0.94 |
| $\sigma_\delta$ | 8.50 | 9.67 | 13.74 | 0.35 | 8.59 | 1.03 | 0.94 | 9.30 | 9.45 | 0.61 |
| Survival |  |  |  |  |  |  |  |  |  |  |
| $b$ | 4.90 | 3.52 | -28.09 | 0.00 | 4.96 | 1.20 | 0.95 | 3.13 | -36.17 | 0.00 |
| $\gamma_0$ | -16.00 | -11.43 | -28.54 | 0.00 | -16.18 | 1.15 | 0.96 | -10.66 | -33.37 | 0.00 |
| $\gamma_1$ | -0.50 | -0.32 | -36.70 | 0.66 | -0.51 | 1.45 | 0.94 | -0.24 | -52.45 | 0.39 |
| $\gamma_2$ | -0.05 | -0.03 | -36.01 | 0.86 | -0.05 | -1.52 | 0.97 | -0.04 | -16.67 | 0.91 |
| $\alpha_1$ | 0.20 | - | - | - | 0.20 | 0.54 | 0.96 | -1.20 | - | - |
| $\alpha_2$ | -0.10 | - | - | - | -0.09 | -7.45 | 0.94 | - | - | - |

True, parameter true value; Mean, posterior mean; PB, percent bias; CP: 95% credible interval; MCDP+RE, the joint model with the shared random effect association structure; MCDP+CV, the joint model with the current value association structure; Separate, the separate models of the MCDP model and the proportional hazards model.

The simulation results (**Table 2**) confirmed that the shared random effect association structure (MCDP+RE) gave rise to the best performance, consistent with our simulating model. In terms of predicted accuracy based on WAIC values, the MCDP+RE model achieved the lowest WAIC (4486 versus 4726 for MCDP+CV and 4993 for MCDP+IND). Both models that addressed informative missingness (MCDP+RE and MCDP+CV) outperformed the model that ignored informative missingness (separate models).

Regarding bias and coverage probability, most of the parameters in the MCDP+RE model showed low bias of smaller than 5%, with coverage probabilities around the nominal rate of 95%. The largest bias of 8% was observed in *l*_2_, the scaling parameter for the exponential curve, but the point estimate (*l*^â^_2_ = −0.0216) was close to the true value of 0.02 with an acceptable coverage probability of 92%. This slightly larger bias was mainly attributed to its relatively small effect size.

Compared to the MCDP+RE model, the joint model with the current value association structure (MCDP+CV) produced larger bias and worse coverage probabilities in the longitudinal sub-model. Specifically, the scaling parameters for the exponential function were overestimated in terms of absolute values (percent bias: 19% for *l*_)_, 6% for *g*_)_, 29% for *l*_2_, and 7% for *g*_2_). The time shift parameters were also overestimated, resulting in 7% bias in the fixed effect β and 9% bias in the standard deviation σ_δ_ of the random effect. The bias indicated larger average differences between individual actual chronological ages and estimated disease ages in the MCDP+CV model versus the MCDP+RE model. The MCDP+CV model incorporated the true unobserved value of longitudinal measures, leading to different survival parameter interpretations compared with the MCDP+RE model. Therefore, the bias and coverage probabilities for the survival parameters in the MCDP+CV model were not available. The simulated true data indicated that participants with a later disease stage (i.e., larger latent time shift) and smaller baseline cognitive measures (i.e., smaller random intercept) had a higher hazard for dropout. The estimated association parameter (αã_=_ = −1.2, 95% CI: −1.37 to −1.03) in the MCDP+CV model indicated that a higher dropout risk was linked to lower current values of longitudinal measures, suggesting participants with these characteristics were likely at a later disease stage and had lower baseline cognitive values. Therefore, despite the misspecified survival sub-model, the MCDP+CV model allows for retrieving the directionality of the association between two processes.

In contrast to the previous two models, the separate models failed to account for informative dropout and yielded the largest bias in the estimated longitudinal parameters (percent bias: 48% for *l*_)_, 11% for *g*_)_, 55% for *l*_2_, 8% for *g*_2_, 10% for β, and 14% for σ_δ_). The overestimated σ_δ_ in the separate models indicated the largest span of latent time shift among the three models.

Figure 4 depicted the mean longitudinal trajectories fitted by three models in two longitudinal measures. The MCDP+RE model produced trajectories that aligned with the true mean trajectories, indicating superior model fit. The estimated trajectories in the separate models displayed the flattest curve, resulting in the greatest deviation from the true trajectories. The underestimated downward trend from the separate models provided a misleadingly optimistic view of disease progression, as it failed to account for informative dropout associated with worsening cognitive conditions. The MCDP+CV model, which addressed informative missingness through a misspecified association structure, produced intermediate estimates. The resulting fitted mean trajectories positioned the MCDP+CV model between the other two models.

**Figure 4.**
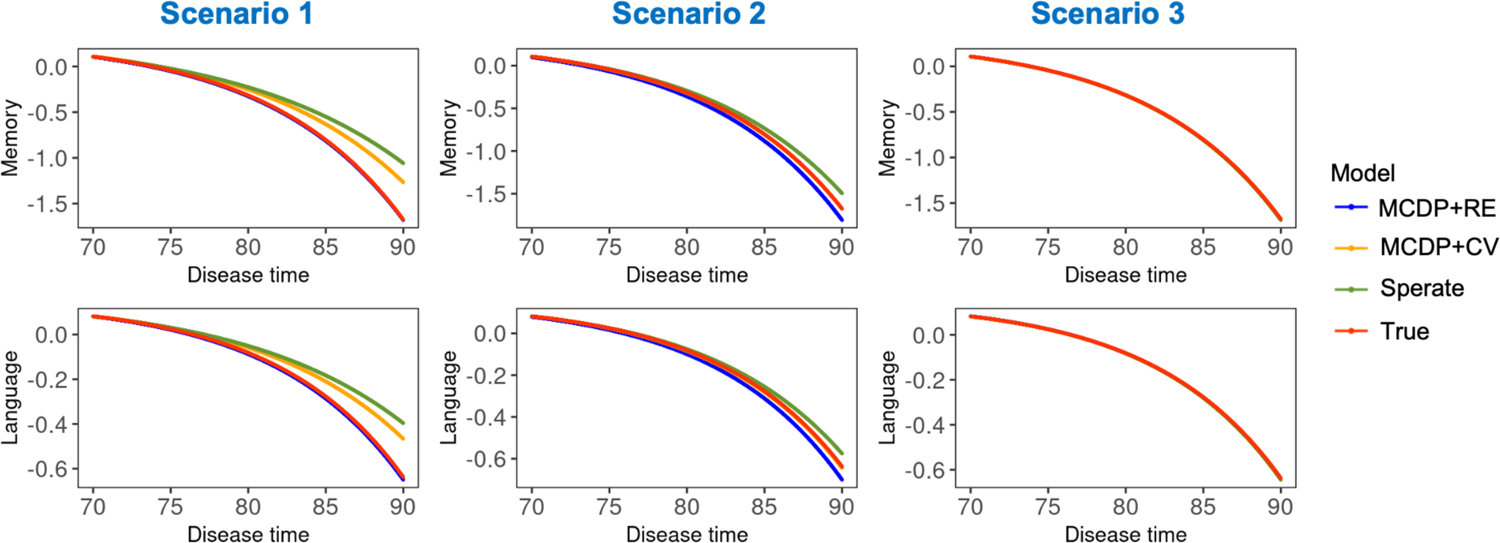
Mean disease progression trajectories fitted by joint models and separate models in Scenarios 1, 2, and 3. The panels in the left, middle and right columns represent the mean disease trajectories in Scenario 1, 2, and 3, respectively. The y-axis in each panel represents the mean values of cognitive measure, and the x-axis denote the disease time estimated from the models. Four curves in each panel show trajectories fitted by different models: MCDP+RE (blue), MCDP+CV (orange), separate models (green), true trajectories (red). Note that if a curve overlaps with the true trajectory, only the red curve representing the true trajectory is displayed. Scenario 1, shared random effect association structure; Scenario 2, current value association structure; Scenario 3, separate (independent) association structure; MCDP+RE, the joint model with the shared random effect association structure; MCDP+CV, the joint model with the current value association structure; Separate, the separate models of the MCDP model and the proportional hazards model.

In order to examine whether our proposed joint models produced more accurate disease stages through latent time shifts, we compared three models using three evaluation metrics: root-mean-square errors (RMSE), median absolute deviations (MAD), and Spearman correlations between estimated and true subject-level latent time shifts across all simulated datasets.

MCDP+RE and the separate models yielded distinct evaluation metrics based on their distributions, while MCDP+CV produced intermediate results (Figure 5). Specifically, the MCDP+RE model yielded the most accurate estimation of latent time shifts with the smallest RMSE and MAD (average RMSE: 4.87 for MCDP+RE versus 5.47 for MCDP+CV and 5.84 for separate models; average MAD: 3 for MCDP+RE versus 3.4 for MCDP+CV and 3.77 for separate models). Besides, the estimated latent time shifts in the MCDP+RE model demonstrated the highest correlations with the true values (average correlation of 0.82 for MCDP+RE versus 0.73 for MCDP+CV and 0.77 for separate models).

**Figure 5.**
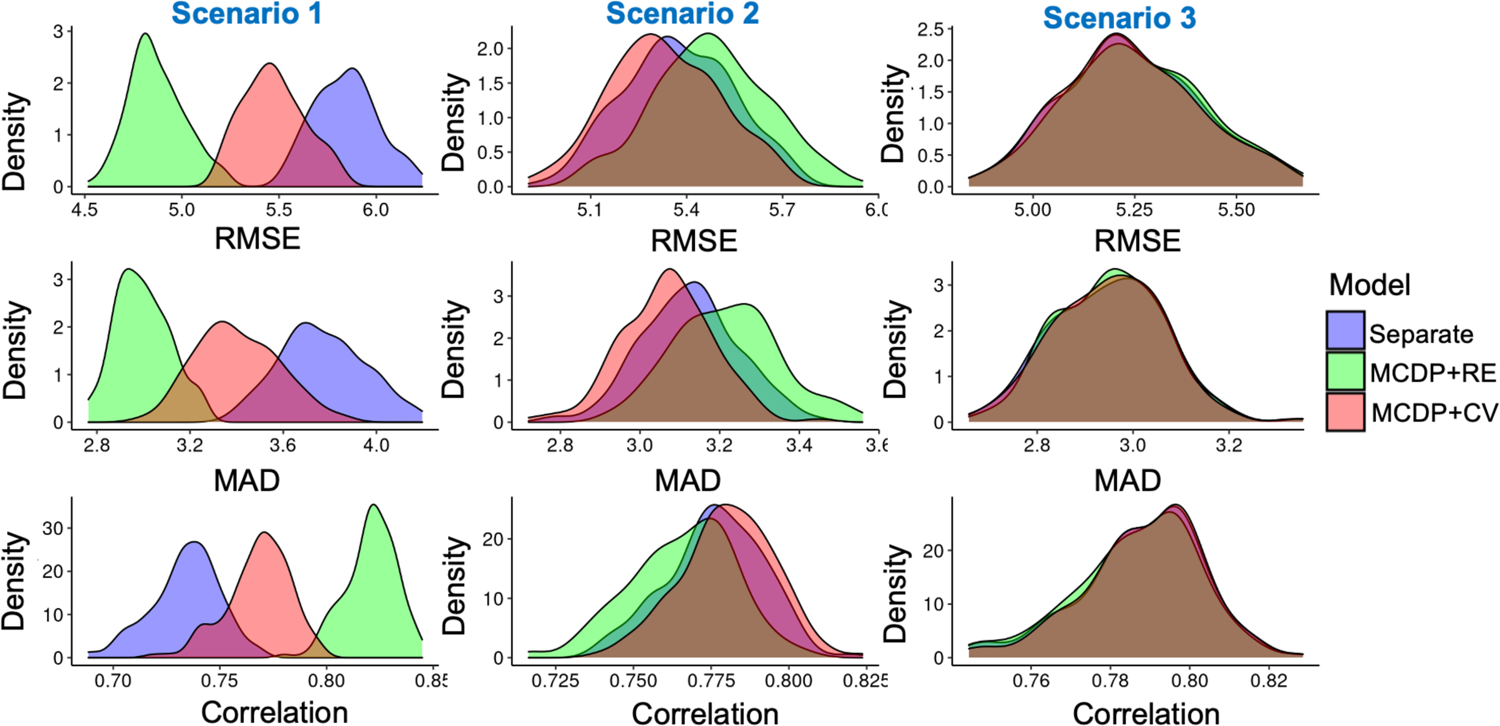
Evaluation metrics of latent time shift for three models in Scenarios 1, 2, and 3. Each of the 9 panels in this figure displays a density plot for one of the evaluation metrics across all simulations. The columns in the figure correspond to different scenarios: the left column shows density plots for Scenario 1, the middle column for Scenario 2, and the right column for Scenario 3. In each scenario, the top, middle, and bottom panels display the distributions for RMSE, MDAE, and correlation, respectively. Scenario 1, shared random effect association structure; Scenario 2, current value association structure; Scenario 3, separate (independent) association structure; MCDP+RE, the joint model with the shared random effect association structure; MCDP+CV, the joint model with the current value association structure; Separate, the separate models of the MCDP model and the proportional hazards model; RMSE, root-mean-square-error; MAD, median absolute deviations.

### Scenario 2: current value data structure (Table 3)

When dropout times were related to the longitudinal process through true unobserved current values of longitudinal measures, the MCDP+CV model achieved the best predictive accuracy based on the smallest WAIC value (4700 versus 4727 for MCDP+RE and 4936 for separate models). The MCDP+RE model also outperformed the separate models. In addition, the MCDP+CV model yielded unbiased estimates, with all model parameters’ bias smaller than 5%.

**Table 3.** Simulation results for Scenario 2 (current value association structure).

| Parameter | True | Separate<br>(WAIC=4936) |  |  | MCDP+RE<br>(WAIC=4727) |  |  | MCDP+CV<br>(WAIC=4700) |  |  |
| --- | --- | --- | --- | --- | --- | --- | --- | --- | --- | --- |
|  |  | Means | Bias | CP | Means | Bias | CP | Means | Bias | CP |
| Longitudinal |  |  |  |  |  |  |  |  |  |  |
| $l_1$ | -0.06 | -0.07 | 12.50 | 0.83 | -0.07 | 12.44 | 0.83 | -0.06 | 3.26 | 0.93 |
| $g_1$ | 2.15 | 2.21 | 2.86 | 0.60 | 2.16 | 0.63 | 0.92 | 2.16 | 0.40 | 0.94 |
| $v_1$ | 0.30 | 0.31 | 2.98 | 0.97 | 0.31 | 3.65 | 0.93 | 0.30 | 0.99 | 0.97 |
| $l_2$ | -0.02 | -0.02 | 15.50 | 0.91 | -0.02 | 12.09 | 0.92 | -0.02 | 4.07 | 0.96 |
| $g_2$ | 2.10 | 2.16 | 2.93 | 0.80 | 2.11 | 0.41 | 0.93 | 2.11 | 0.37 | 0.94 |
| $v_2$ | 0.15 | 0.16 | 3.66 | 0.93 | 0.15 | 3.14 | 0.92 | 0.15 | 1.53 | 0.94 |
| $\sigma_1$ | 0.30 | 0.30 | 0.40 | 0.92 | 0.30 | 0.30 | 0.93 | 0.30 | 0.09 | 0.94 |
| $\sigma_2$ | 0.20 | 0.20 | 0.04 | 0.99 | 0.20 | 0.05 | 0.99 | 0.20 | 0.01 | 0.99 |
| $\sigma_{b1}$ | 0.45 | 0.45 | 0.10 | 0.93 | 0.45 | -0.47 | 0.93 | 0.45 | -0.24 | 0.92 |
| $\sigma_{b2}$ | 0.50 | 0.50 | -0.27 | 0.94 | 0.50 | -0.44 | 0.94 | 0.50 | -0.46 | 0.94 |
| $\rho_b$ | 0.60 | 0.60 | -0.03 | 0.93 | 0.60 | -0.38 | 0.93 | 0.60 | -0.31 | 0.92 |
| Time shift |  |  |  |  |  |  |  |  |  |  |
| $\beta$ | -2.00 | -2.04 | 2.18 | 0.95 | -1.98 | -0.84 | 0.96 | -2.02 | 1.11 | 0.94 |
| $\sigma_\delta$ | 8.50 | 8.86 | 4.24 | 0.85 | 8.30 | -2.39 | 0.91 | 8.60 | 1.19 | 0.94 |
| Survival |  |  |  |  |  |  |  |  |  |  |
| $b$ | 2.90 | 3.44 | 18.58 | 0.16 | 3.92 | 35.10 | 0.00 | 2.94 | 1.27 | 0.93 |
| $\gamma_0$ | -10.00 | -11.19 | 11.91 | 0.39 | -12.83 | 28.28 | 0.00 | -10.13 | 1.31 | 0.93 |
| $\gamma_1$ | -0.20 | -0.29 | 45.49 | 0.85 | -0.36 | 79.00 | 0.77 | -0.20 | -1.86 | 0.94 |
| $\gamma_2$ | -0.05 | -0.04 | -19.98 | 0.91 | -0.04 | -11.06 | 0.96 | -0.05 | -4.58 | 0.97 |
| $\alpha_1$ | -1.00 | - | - | - | 0.11 | - | - | -1.01 | 0.74 | 0.97 |
| $\alpha_2$ | - | - | - | - | -0.84 | - | - | - | - | - |

True, parameter true value; Mean, posterior mean; PB, percent bias; CP: 95% credible interval; MCDP+RE, the joint model with the shared random effect association structure; MCDP+CV, the joint model with the current value association structure; Separate, the separate models of the MCDP model and the proportional hazards model.

Misspecifying the association structure using the MCDP+RE model slightly biased the longitudinal parameter estimates (percent bias: 12% for *l*_)_ and 12% for *l*_2_). Notably, the estimated span of latent time shift was slightly smaller than that in the MCDP+CV model (8.3 versus 8.6). Despite the misspecified association structure, the direction of the association between two processes was captured by the MCDP+RE model. The association parameters (αã_=_ = 0.11, αã_>_ = −0.84) indicated that a larger latent time shift and a smaller random intercept in the cognitive measure—both associated with lower cognitive values—were correlated with an increased risk of dropout. This finding aligned with the negative association between the dropout times and longitudinal measures in the true data.

Like in Scenario 1, the separate models generated the most biased estimates in the longitudinal measures (percent bias: 13% for *l*_)_, 16% for *l*_2_) with an overestimated deviation of latent time shifts (4% bias for σ_δ_).

In Figure 4, the estimated mean cognitive trajectories from the MCDP+CV model overlapped with the true trajectories. In contrast, the MCDP+RE model produced slightly steeper curves, indicating an overcorrection for informative dropout. The separate models generated trajectories with a slower acceleration in their rate of decline compared to the joint models, providing overoptimistic estimates. In Figure 5 for the evaluation of latent time shifts, most parts of the distributions overlapped, suggesting only minor differences among the models.

Nevertheless, the latent time shifts estimated from the MCDP+CV model were the most accurate (average RMSE: 5.33 versus 5.37 and 5.48; average MAD: 3.07 versus 3.12 and 3.22; average correlation: 0.78 versus 0.78 and 0.77).

### Scenario 3: separate data structure (Table 4)

When dropouts were not informative, WAIC values were comparable across all three models (4661 for separate models, 4661 for MCDP+RE, and 4658 for MCDP+CV). All three models produced unbiased estimates, with percent bias smaller than 2%. Notably, the association parameters in the models that treat dropouts as informative were close to 0. In addition, the mean trajectories estimated by all three models closely matched the true trajectories, as shown in Figure 4. Furthermore, the evaluation metrics for the latent time shift were nearly identical across the three models (Figure 5). These findings suggested that all models performed well, irrespective of assumptions about informative missingness.

**Table 4.** Simulation results for Scenario 3 (separate data structure).

| Parameter | True | Separate<br>(WAIC=4661) |  |  | MCDP+RE<br>(WAIC=4661) |  |  | MCDP+CV<br>(WAIC=4658) |  |  |
| --- | --- | --- | --- | --- | --- | --- | --- | --- | --- | --- |
|  |  | Means | Bias | CP | Means | Bias | CP | Means | Bias | CP |
| Longitudinal |  |  |  |  |  |  |  |  |  |  |
| $l_1$ | -0.06 | -0.06 | 0.52 | 0.95 | -0.06 | 0.04 | 0.95 | -0.06 | 0.16 | 0.96 |
| $g_1$ | 2.15 | 2.15 | 0.00 | 0.93 | 2.15 | -0.02 | 0.93 | 2.15 | -0.04 | 0.94 |
| $v_1$ | 0.30 | 0.30 | 0.00 | 0.97 | 0.30 | -0.12 | 0.97 | 0.30 | -0.01 | 0.97 |
| $l_2$ | -0.02 | -0.02 | 1.23 | 0.95 | -0.02 | 0.96 | 0.98 | -0.02 | 1.10 | 0.96 |
| $g_2$ | 2.10 | 2.10 | 0.03 | 0.94 | 2.10 | 0.04 | 0.95 | 2.10 | 0.01 | 0.95 |
| $v_2$ | 0.15 | 0.15 | 0.79 | 0.93 | 0.15 | 1.13 | 0.93 | 0.15 | 0.87 | 0.94 |
| $\sigma_1$ | 0.30 | 0.30 | 0.10 | 0.94 | 0.30 | 0.10 | 0.93 | 0.30 | 0.09 | 0.94 |
| $\sigma_2$ | 0.20 | 0.20 | 0.01 | 0.98 | 0.20 | 0.00 | 0.98 | 0.20 | -0.02 | 0.98 |
| $\sigma_{b1}$ | 0.45 | 0.45 | -0.12 | 0.93 | 0.45 | -0.18 | 0.93 | 0.45 | -0.14 | 0.93 |
| $\sigma_{b2}$ | 0.50 | 0.50 | -0.34 | 0.95 | 0.50 | -0.37 | 0.94 | 0.50 | -0.30 | 0.93 |
| $\rho_b$ | 0.60 | 0.60 | -0.17 | 0.93 | 0.60 | -0.16 | 0.95 | 0.60 | -0.12 | 0.94 |
| Time shift |  |  |  |  |  |  |  |  |  |  |
| $\beta$ | -2.00 | -2.03 | 1.30 | 0.97 | -2.01 | 0.49 | 0.95 | -2.01 | 0.58 | 0.96 |
| $\sigma_\delta$ | 8.50 | 8.53 | 0.41 | 0.95 | 8.54 | 0.43 | 0.95 | 8.55 | 0.55 | 0.96 |
| Survival |  |  |  |  |  |  |  |  |  |  |
| $b$ | 3.70 | 3.72 | 0.53 | 0.90 | 3.73 | 0.80 | 0.90 | 3.73 | 0.72 | 0.92 |
| $\gamma_0$ | -12.00 | -12.07 | 0.58 | 0.90 | -12.11 | 0.89 | 0.91 | -12.09 | 0.73 | 0.91 |
| $\gamma_1$ | -0.50 | -0.51 | 1.11 | 0.93 | -0.51 | 1.79 | 0.93 | -0.51 | 1.20 | 0.94 |
| $\gamma_2$ | -0.10 | -0.10 | -1.21 | 0.99 | -0.10 | -0.95 | 0.98 | -0.10 | -1.25 | 0.99 |
| $\alpha_1$ | - | - | - | - | 0.00 | - | - | 0.02 | - | - |
| $\alpha_2$ | - | - | - | - | 0.00 | - | - | - | - | - |

True, parameter true value; Mean, posterior mean; PB, percent bias; CP: 95% credible interval; MCDP+RE, the joint model with the shared random effect association structure; MCDP+CV, the joint model with the current value association structure; Separate, the separate models of the MCDP model and the proportional hazards model.

### Other scenarios

Scenarios 4, 5, and 6 adopted the shared random effect data structure as used in Scenario 1 but had smaller sample size (n=500 versus 1000 in Scenario 4), deviation of the random latent time shift (σ_δ_ = 4 versus σ_δ_ = 8.5 in Scenario 5), and event rate (50% versus 30% in Scenario 6).

According to **Tables 5**, **6**, and **7**, reducing sample size, limiting the span of latent time shift, or increasing event rate caused more biased estimates for all three models compared with Scenario 1. The conclusions regarding model comparison from Scenario 1 also applied to Scenarios 4, 5, and 6. Notably, lowering the span of latent time shift minimized the difference between the joint models with misspecified structure (MCDP+CV) and the separate models (Figures 6 **and 7**). In addition, a larger proportion of informative dropout more clearly differentiate the performance of the correctly specified model (MCDP+RE) from the other two models (Figures 6 **and 7**), with the best performance from MCDP+RE.

**Figure 6.**
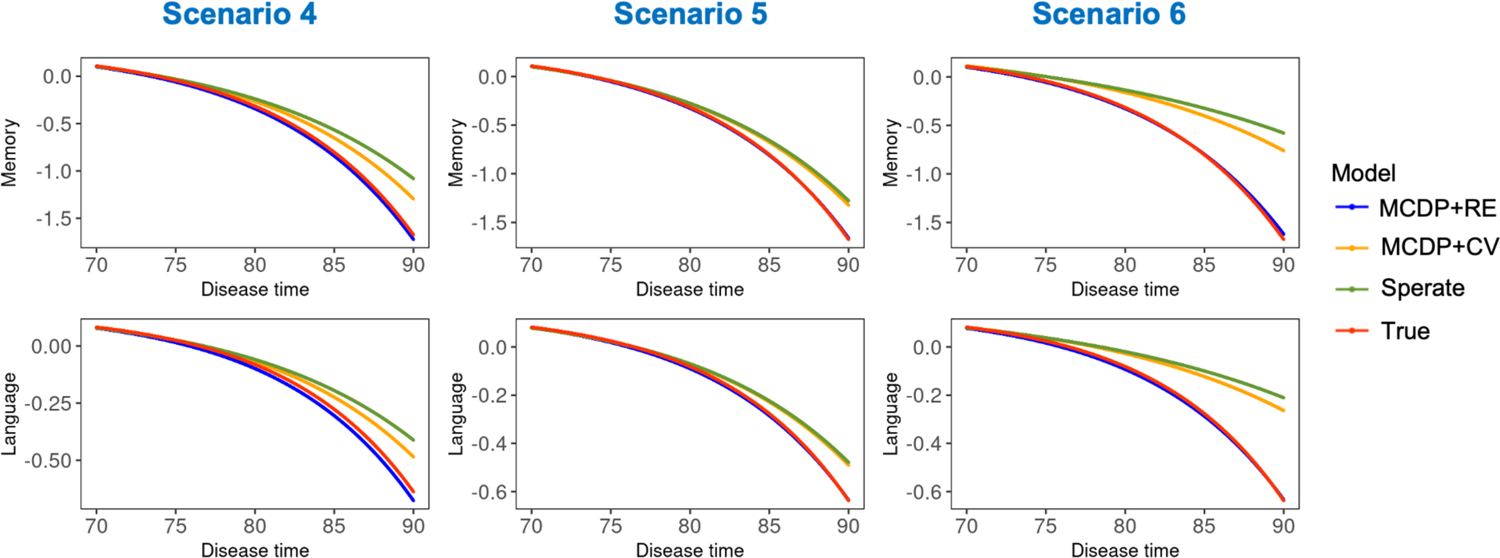
Mean disease progression trajectories fitted by joint models and separate models in Scenarios 4, 5, and 6. The panels in the left, middle and right columns represent the mean disease trajectories in Scenario 1, 2, and 3, respectively. The y-axis in each panel represents the mean values of cognitive measure, and the x-axis denote the disease time estimated from the models. Four curves in each panel show trajectories fitted by different models: MCDP+RE (blue), MCDP+CV (orange), separate models (green), true trajectories (red). Note that if a curve overlaps with the true trajectory, only the red curve representing the true trajectory is displayed. Scenario 4, shared random effect association structure with N=500; Scenario 5, shared random effect association structure with σ_δ_ = 4; Scenario 6, shared random effect association structure with 50% event rate; MCDP+RE, the joint model with the shared random effect association structure; MCDP+CV, the joint model with the current value association structure; Separate, the separate models of the MCDP model and the proportional hazards model.

**Figure 7.**
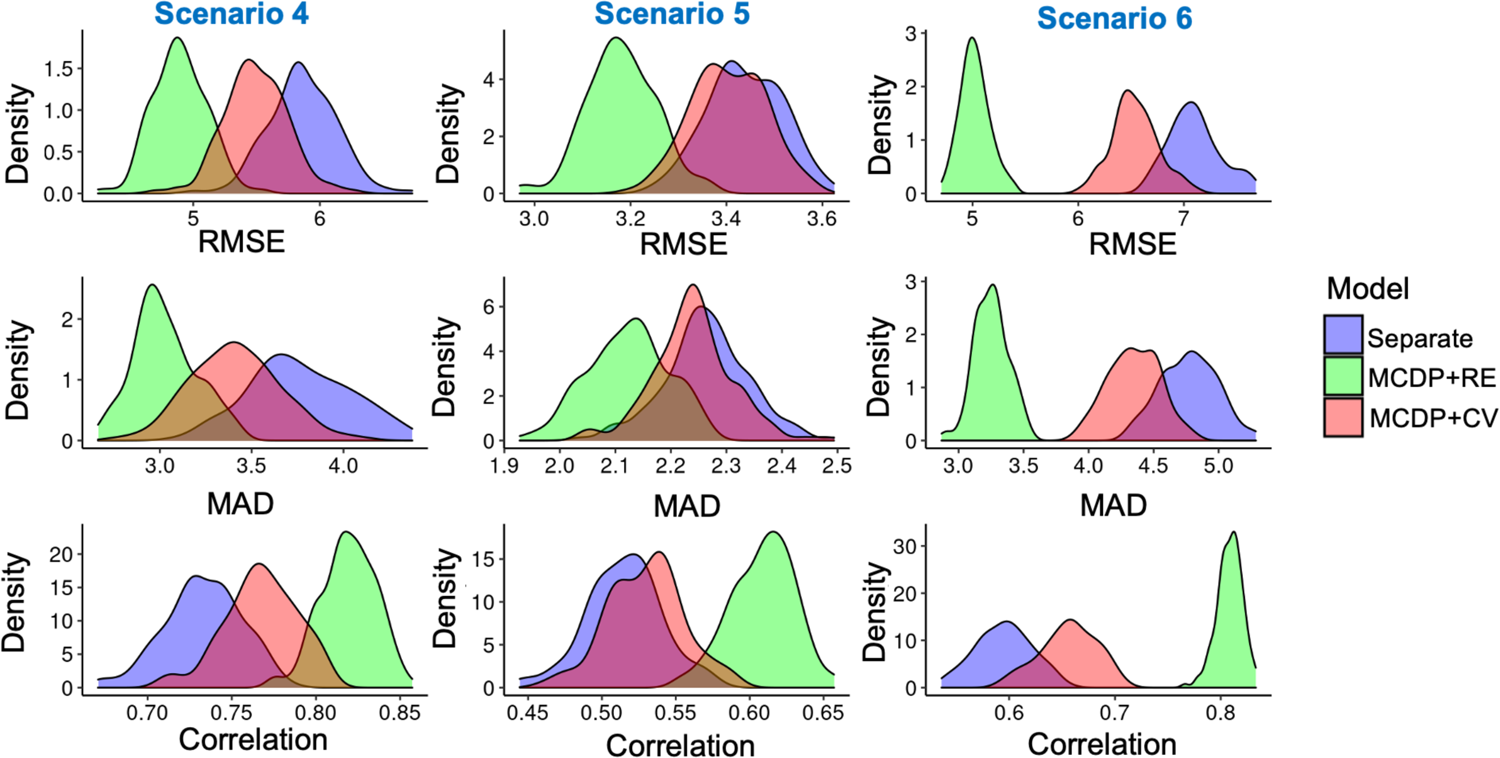
Evaluation metrics of latent time shift for three models in Scenarios 4, 5, and 6. Each of the 9 panels in this figure displays a density plot for one of the evaluation metrics across all simulations. The columns in the figure correspond to different scenarios: the left column shows density plots for Scenario 4, the middle column for Scenario 5, and the right column for Scenario 6. In each scenario, the top, middle, and bottom panels display the distributions for RMSE, MDAE, and correlation, respectively. Scenario 4, shared random effect association structure with N=500; Scenario 5, shared random effect association structure with σ_δ_ = 4; Scenario 6, shared random effect association structure with 50% event rate; MCDP+RE, the joint model with the shared random effect association structure; MCDP+CV, the joint model with the current value association structure; Separate, the separate models of the MCDP model and the proportional hazards model; RMSE, root-mean-square-error; MAD, median absolute deviations.

**Table 5.** Simulation results for Scenario 4 (shared random effect structure with N=500).

| Parameter | True | Separate<br>(WAIC=2497) |  |  | MCDP+RE<br>(WAIC=2232) |  |  | MCDP+CV<br>(WAIC=2367) |  |  |
| --- | --- | --- | --- | --- | --- | --- | --- | --- | --- | --- |
|  |  | Means | Bias | CP | Means | Bias | CP | Means | Bias | CP |
| Longitudinal |  |  |  |  |  |  |  |  |  |  |
| $l_1$ | -0.06 | -0.09 | 56.05 | 0.41 | -0.07 | 11.41 | 0.95 | -0.08 | 25.56 | 0.88 |
| $g_1$ | 2.15 | 2.40 | 11.74 | 0.06 | 2.17 | 1.08 | 0.94 | 2.28 | 6.14 | 0.56 |
| $v_1$ | 0.30 | 0.33 | 10.71 | 0.81 | 0.31 | 2.60 | 0.96 | 0.31 | 4.74 | 0.92 |
| $l_2$ | -0.02 | -0.03 | 73.04 | 0.57 | -0.02 | 19.82 | 0.95 | -0.03 | 43.88 | 0.83 |
| $g_2$ | 2.10 | 2.37 | 12.64 | 0.20 | 2.13 | 1.65 | 0.93 | 2.27 | 7.90 | 0.58 |
| $v_2$ | 0.15 | 0.17 | 11.32 | 0.93 | 0.16 | 4.03 | 0.96 | 0.16 | 7.20 | 0.95 |
| $\sigma_1$ | 0.30 | 0.30 | 0.98 | 0.85 | 0.30 | 0.16 | 0.94 | 0.30 | 0.30 | 0.92 |
| $\sigma_2$ | 0.20 | 0.20 | 0.17 | 0.98 | 0.20 | -0.04 | 0.98 | 0.20 | -0.02 | 0.98 |
| $\sigma_{b1}$ | 0.45 | 0.45 | -0.14 | 0.91 | 0.45 | -0.41 | 0.91 | 0.45 | -0.13 | 0.92 |
| $\sigma_{b2}$ | 0.50 | 0.50 | -0.55 | 0.95 | 0.50 | -0.64 | 0.92 | 0.50 | -0.53 | 0.94 |
| $\rho_b$ | 0.60 | 0.60 | -0.42 | 0.93 | 0.60 | -0.73 | 0.93 | 0.60 | -0.53 | 0.94 |
| Time shift |  |  |  |  |  |  |  |  |  |  |
| $\beta$ | -2.00 | -2.30 | 15.12 | 0.93 | -2.20 | 9.75 | 0.94 | -2.28 | 13.98 | 0.94 |
| $\sigma_\delta$ | 8.50 | 9.70 | 14.15 | 0.63 | 8.62 | 1.43 | 0.92 | 9.37 | 10.24 | 0.76 |
| Survival |  |  |  |  |  |  |  |  |  |  |
| $b$ | 4.90 | 3.52 | -28.15 | 0.00 | 5.03 | 2.58 | 0.96 | 3.12 | -36.23 | 0.00 |
| $\gamma_0$ | -16.00 | -11.43 | -28.58 | 0.00 | -16.40 | 2.50 | 0.95 | -10.66 | -33.35 | 0.00 |
| $\gamma_1$ | -0.50 | -0.33 | -34.24 | 0.85 | -0.54 | 8.36 | 0.93 | -0.24 | -51.21 | 0.63 |
| $\gamma_2$ | -0.05 | -0.03 | -31.28 | 0.89 | -0.05 | 3.72 | 0.95 | -0.04 | -13.51 | 0.93 |
| $\alpha_1$ | 0.20 | - | - | - | 0.21 | 3.44 | 0.95 | -1.22 | - | - |
| $\alpha_2$ | -0.10 | - | - | - | -0.07 | -30.13 | 0.96 | - | - | - |

True, parameter true value; Mean, posterior mean; PB, percent bias; CP: 95% credible interval; MCDP+RE, the joint model with the shared random effect association structure; MCDP+CV, the joint model with the current value association structure; Separate, the separate models of the MCDP model and the proportional hazards model.

**Table 6.** Simulation results for Scenario 5 (shared random effect structure with σ_δ_=4).

| Parameter | True | Separate<br>(WAIC=4529) |  |  | MCDP+RE<br>(WAIC=4430) |  |  | MCDP+CV<br>(WAIC=4510) |  |  |
| --- | --- | --- | --- | --- | --- | --- | --- | --- | --- | --- |
|  |  | Means | Bias | CP | Means | Bias | CP | Means | Bias | CP |
| Longitudinal |  |  |  |  |  |  |  |  |  |  |
| $l_1$ | -0.06 | -0.09 | 42.77 | 0.50 | -0.07 | 8.98 | 0.94 | -0.08 | 37.17 | 0.59 |
| $g_1$ | 2.15 | 2.33 | 8.18 | 0.20 | 2.18 | 1.21 | 0.95 | 2.30 | 7.14 | 0.32 |
| $v_1$ | 0.30 | 0.33 | 9.33 | 0.80 | 0.31 | 2.07 | 0.95 | 0.32 | 8.10 | 0.84 |
| $l_2$ | -0.02 | -0.03 | 53.46 | 0.71 | -0.02 | 17.83 | 0.96 | -0.03 | 49.59 | 0.77 |
| $g_2$ | 2.10 | 2.29 | 9.00 | 0.46 | 2.14 | 2.12 | 0.95 | 2.28 | 8.35 | 0.53 |
| $v_2$ | 0.15 | 0.16 | 8.79 | 0.91 | 0.16 | 3.51 | 0.93 | 0.16 | 8.06 | 0.90 |
| $\sigma_1$ | 0.30 | 0.30 | 0.46 | 0.94 | 0.30 | 0.19 | 0.95 | 0.30 | 0.35 | 0.94 |
| $\sigma_2$ | 0.20 | 0.20 | 0.12 | 0.98 | 0.20 | 0.05 | 0.97 | 0.20 | 0.10 | 0.98 |
| $\sigma_{b1}$ | 0.45 | 0.45 | -0.32 | 0.93 | 0.45 | -0.18 | 0.94 | 0.45 | -0.36 | 0.93 |
| $\sigma_{b2}$ | 0.50 | 0.50 | -0.34 | 0.93 | 0.50 | -0.30 | 0.93 | 0.50 | -0.29 | 0.94 |
| $\rho_b$ | 0.60 | 0.60 | -0.08 | 0.93 | 0.60 | -0.09 | 0.93 | 0.60 | -0.07 | 0.92 |
| Time shift |  |  |  |  |  |  |  |  |  |  |
| $\beta$ | -2.00 | -2.09 | 4.74 | 0.93 | -2.08 | 4.21 | 0.97 | -2.11 | 5.34 | 0.93 |
| $\sigma_\delta$ | 4.00 | 4.24 | 5.93 | 0.89 | 4.04 | 0.97 | 0.97 | 4.25 | 6.23 | 0.89 |
| Survival |  |  |  |  |  |  |  |  |  |  |
| $b$ | 5.00 | 4.57 | -8.50 | 0.51 | 5.07 | 1.38 | 0.94 | 4.35 | -13.08 | 0.22 |
| $\gamma_0$ | -16.00 | -14.60 | -8.75 | 0.44 | -16.22 | 1.37 | 0.94 | -14.00 | -12.49 | 0.21 |
| $\gamma_1$ | -0.50 | -0.45 | -10.99 | 0.93 | -0.52 | 3.68 | 0.94 | -0.41 | -17.82 | 0.88 |
| $\gamma_2$ | -0.05 | -0.04 | -14.28 | 0.97 | -0.05 | -1.70 | 0.97 | -0.04 | -11.74 | 0.96 |
| $\alpha_1$ | 0.20 | - | - | - | 0.20 | 1.38 | 0.95 | -0.43 | - | - |
| $\alpha_2$ | -0.10 | - | - | - | -0.11 | 12.42 | 0.95 | - | - | - |

True, parameter true value; Mean, posterior mean; PB, percent bias; CP: 95% credible interval; MCDP+RE, the joint model with the shared random effect association structure; MCDP+CV, the joint model with the current value association structure; Separate, the separate models of the MCDP model and the proportional hazards model.

**Table 7.** Simulation results for Scenario 6 (shared random effect structure with 50% event rate).

| Parameter | True | Separate<br>(WAIC=6190) |  |  | MCDP+RE<br>(WAIC=5502) |  |  | MCDP+CV<br>(WAIC=5993) |  |  |
| --- | --- | --- | --- | --- | --- | --- | --- | --- | --- | --- |
|  |  | Means | Bias | CP | Means | Bias | CP | Means | Bias | CP |
| Longitudinal |  |  |  |  |  |  |  |  |  |  |
| $l_1$ | -0.06 | -0.15 | 156.57 | 0.02 | -0.07 | 15.43 | 0.91 | -0.10 | 72.75 | 0.40 |
| $g_1$ | 2.15 | 2.79 | 29.89 | 0.00 | 2.20 | 2.26 | 0.88 | 2.54 | 18.21 | 0.02 |
| $v_1$ | 0.30 | 0.39 | 29.33 | 0.34 | 0.31 | 3.42 | 0.96 | 0.34 | 13.21 | 0.72 |
| $l_2$ | -0.02 | -0.06 | 179.85 | 0.13 | -0.02 | 24.02 | 0.94 | -0.04 | 110.27 | 0.43 |
| $g_2$ | 2.10 | 2.73 | 29.87 | 0.00 | 2.16 | 2.82 | 0.93 | 2.55 | 21.46 | 0.03 |
| $v_2$ | 0.15 | 0.19 | 24.80 | 0.62 | 0.16 | 4.39 | 0.94 | 0.17 | 15.19 | 0.82 |
| $\sigma_1$ | 0.30 | 0.31 | 1.77 | 0.69 | 0.30 | 0.24 | 0.92 | 0.30 | 0.66 | 0.87 |
| $\sigma_2$ | 0.20 | 0.20 | 0.36 | 0.97 | 0.20 | -0.02 | 0.96 | 0.20 | 0.16 | 0.96 |
| $\sigma_{b1}$ | 0.45 | 0.44 | -1.68 | 0.89 | 0.45 | -0.41 | 0.93 | 0.45 | -1.10 | 0.92 |
| $\sigma_{b2}$ | 0.50 | 0.50 | -0.57 | 0.95 | 0.50 | -0.43 | 0.93 | 0.50 | -0.36 | 0.95 |
| $\rho_b$ | 0.60 | 0.60 | -0.02 | 0.93 | 0.60 | -0.36 | 0.93 | 0.60 | -0.02 | 0.92 |
| Time shift |  |  |  |  |  |  |  |  |  |  |
| $\beta$ | -2.00 | -2.58 | 28.95 | 0.95 | -2.12 | 5.82 | 0.95 | -2.45 | 22.25 | 0.94 |
| $\sigma_\delta$ | 8.50 | 11.77 | 38.51 | 0.03 | 8.70 | 2.40 | 0.92 | 10.93 | 28.54 | 0.05 |
| Survival |  |  |  |  |  |  |  |  |  |  |
| $b$ | 5.00 | 3.21 | -35.86 | 0.00 | 5.09 | 1.89 | 0.93 | 3.09 | -38.26 | 0.00 |
| $\gamma_0$ | -15.00 | -9.72 | -35.20 | 0.00 | -15.28 | 1.86 | 0.92 | -9.48 | -36.81 | 0.00 |
| $\gamma_1$ | -0.50 | -0.29 | -41.92 | 0.33 | -0.52 | 3.44 | 0.94 | -0.25 | -49.52 | 0.22 |
| $\gamma_2$ | -0.05 | -0.03 | -42.35 | 0.68 | -0.05 | 2.59 | 0.95 | -0.03 | -32.89 | 0.77 |
| $\alpha_1$ | 0.20 | NA | NA | NA | 0.20 | 0.96 | 0.97 | -0.96 | NA | NA |
| $\alpha_2$ | -0.10 | NA | NA | NA | -0.08 | -24.94 | 0.97 | NA | NA | NA |

True, parameter true value; Mean, posterior mean; PB, percent bias; CP: 95% credible interval; MCDP+RE, the joint model with the shared random effect association structure; MCDP+CV, the joint model with the current value association structure; Separate, the separate models of the MCDP model and the proportional hazards model.

## 5. Real data application

In this section, we used longitudinal data from neuropsychological tests (NP tests) along with dementia and death data in the Framingham Heart Study (FHS) Offspring cohort to demonstrate our methods. Specifically, we applied our proposed joint models and the separate models to the FHS data and compared their model performance using WAIC and mean disease progression.

### 5.1 Data description

The FHS is a multigenerational cohort study initiated in 1948 by enrolling 5209 residents from Framingham, Massachusetts, in the Original cohort [44]. In 1971, 5214 participants who were offspring of the Original cohort and the spouses of these offspring were included in the Offspring cohort [45]. The participants in the Offspring cohort have undergone up to 10 examinations, which were scheduled every four to six years. Beginning in 1999, the surviving participants in the Offspring cohort were invited to join an ancillary study, where they were administered a battery of neuropsychological (NP) tests every five or six years [2]. Participants identified as having potential cognitive impairment were invited to undergo additional, annual neurologic and NP tests. A dementia review panel with at least one neurologist and one neuropsychologist determined whether the participants had dementia, as well as the dementia type and the date of onset by reviewing every case of possible cognitive decline based on the participants’ cognitive information such as neurologic and NP tests, medical records, and neuroimaging studies [2, 46].

NP tests are used to measure participants’ cognitive changes over time. The NP test measures cover four domains in the FHS: memory, attention and executive function, visuoperceptual, and language [47]. In this research project, we mainly focus on memory and language domains. The memory domain includes the following tests: Wechsler Memory Scale (WMS) Logical Memory – Immediate Recall, WMS Logical Memory – Delayed Recall, WMS Visual Reproductions – Immediate Recall, WMS Visual Reproductions – Delayed Recall, WMS Paired Associates – Immediate Recall, and WMS Paired Associates – Delayed Recall. The language domain includes Boston Naming Test 30 item version, Wide Range Achievement Test-3 (WRAT-3) Reading subtest, and Wechsler Adult Intelligence Scale (WAIS) Similarities subtest. For each participant at each visit, we computed a memory score by averaging the individual’s Z scores from NP tests within the memory domain. Similarly, we calculated a language score by averaging the Z scores from NP tests in the attention and executive function domain.(Figure 9).

Of the 5124 Offspring cohort participants, 2992 had at least one NP test in the memory domain, while 2971 had at least one NP test in the language domain. We excluded participants with missing education information from our analysis: 471 out of 2992 participants for the memory domain, and 466 out of 2971 participants for the language domain. In addition, we restricted the data to participants who had at least three visits with NP tests in both memory and language domains, leaving 1128 participants. Of the 1128 participants, we excluded 11 participants who had dementia onset before the age of 60 at baseline. Our final sample included 1117 participants with 4162 observations. Of the 1117 participants in our final sample, 86 were diagnosed with dementia and 231 died during follow-up.

### 5.2 Model specification

In this section, we consider three scenarios regarding informative dropout in the survival sub-model: (i) dropout due to dementia only (event rate: 7.7%); (ii) dropout due to death only (event rate: 20.7%); (iii) dropout due to either dementia or death (combined)(event rate: 28.4%).

Competing risks and semi-competing risks are beyond the scope of this chapter and will be further investigated in depth in the next chapter. In the longitudinal part of the joint model, we fitted the MCDP model for the z-scores in the memory and language domains. The timescale is age that is centered at 60 years. Covariates include sex and education in years. We assume that two cognitive measures, i.e., memory z-scores and language z-scores, are linked through the latent time shift and the random intercepts. The MCDP model is shown as follows:

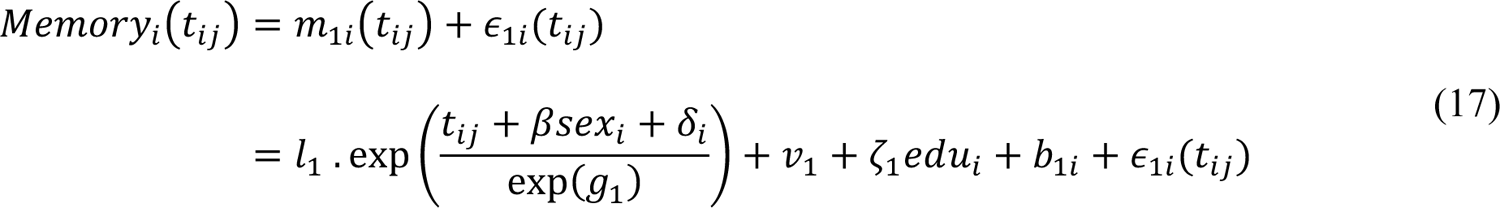

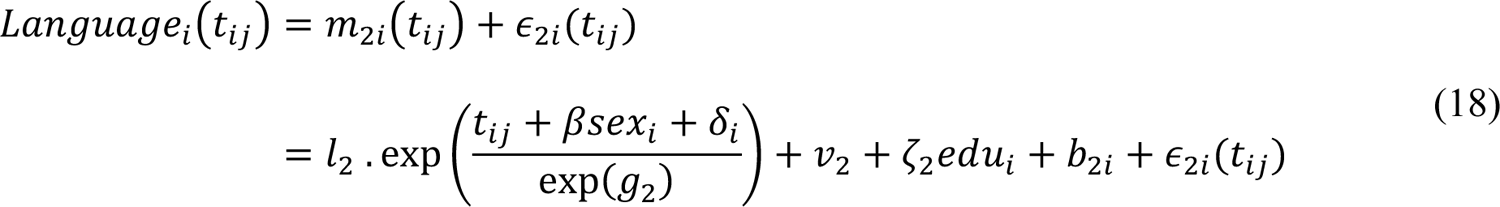

where β is the fixed effect of sex, which is assumed to have an impact on the estimation of the latent time shift. δ_i_ is the random part in the latent time shift that follows a normal distribution: δ_i_∼*N*(0, σ^2^). Education has an influence on the mean cognitive levels with coefficients ॐ_1_ and ॐ_2_ for the memory and language z-scores, respectively. *b*_1i_ and *b*_2i_ are random intercepts for the memory and language z-scores, respectively, that follow a multivariate normal distribution: 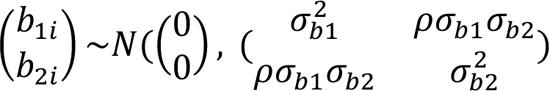

The models are constructed by combining the MCDP model with either of the following three survival sub-models. Time in the survival sub-model is age at the occurrence of the event of our interest (i.e., death, dementia, or the composite survival outcome for both death and dementia) subtracted by 60 years to be consistent with the timescale in the longitudinal part. The following sub-models with three different association structures are considered:

### Shared random effect association structure (RE)

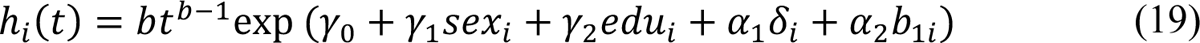

### Current value association structure (CV)

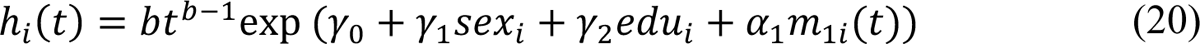

### Separate models

Both sex and education are included as covariates. All other settings are the same as in our simulation study. Convergence was assessed using the potential scale reduction statistic Ř [40]. Models were compared based on their WAIC values.

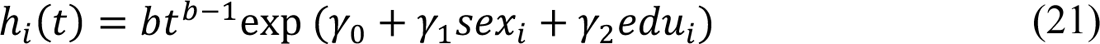

### 5.3 Results

We applied latent time shifts in our proposed joint models and the separate models and observed that trajectories for cognitive measures were re-aligned along a new global disease timeline (Figure 8). For the memory domain, the realigned mean disease progression trajectories showed a more pronounced downward trend compared with those on the original timescale. The modification in the mean disease progression trajectories for the language domain was less distinct, but the slight upward trend observed in the original data was mitigated after the application of latent time shifts.

**Figure 8.**
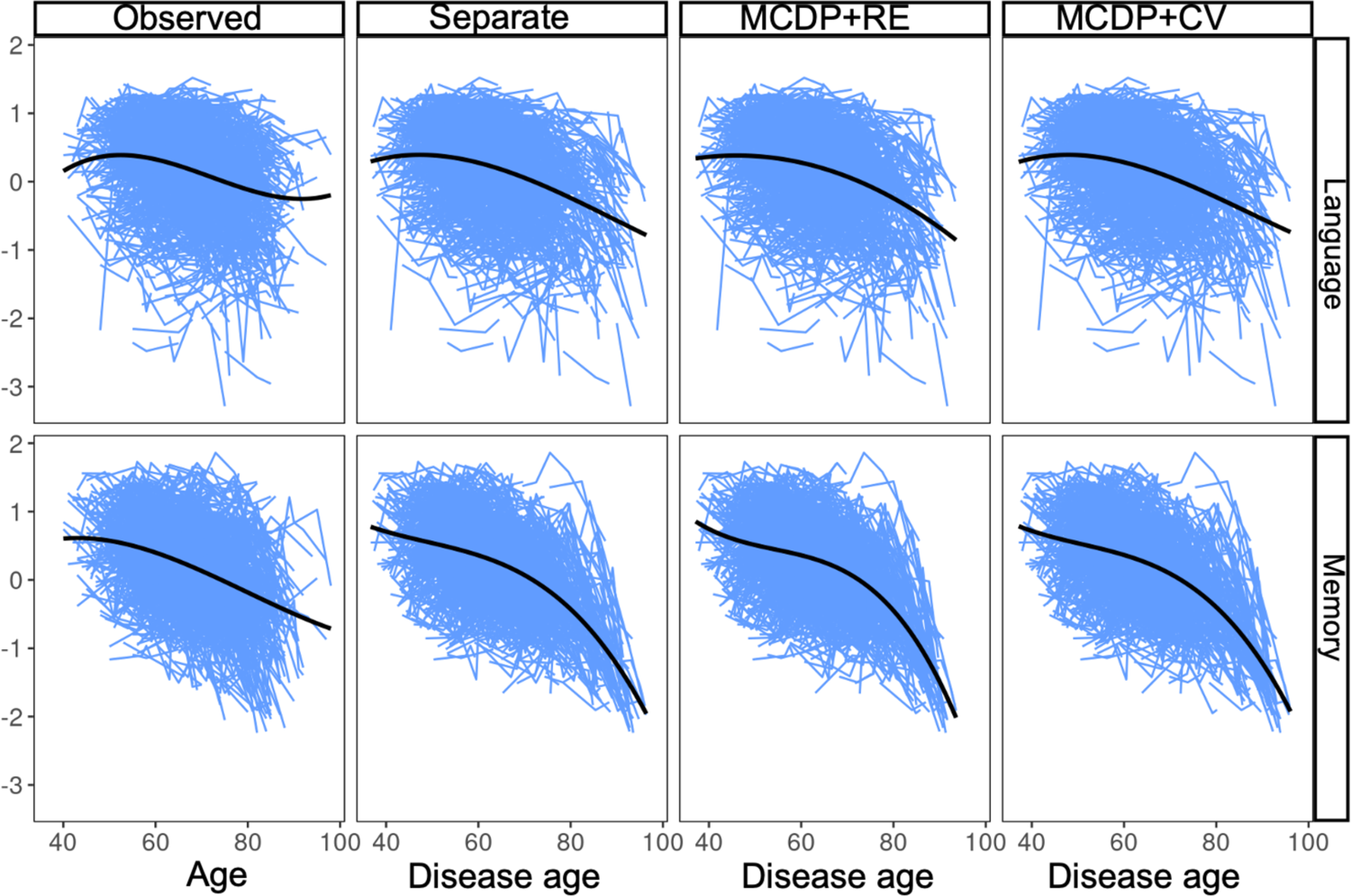
Cognitive trajectories on the original time and disease time in the FHS. The eight panels illustrate the trajectories for the language and memory z-scores in the FHS data. The y-axis in each panel denotes the value of cognitive measures, and the x-axis represents actual chronological age (first column) and disease age (other columns). The two panels in the first column denotes the trajectories aligned on the original actual chronological age. The other panels represent the trajectories that are realigned by fitting different models with latent time shifts. MCDP+RE, the joint model with the shared random effect association structure; MCDP+CV, the joint model with the current value association structure; Separate, the separate models of the MCDP model and the proportional hazards model; FHS, Framingham Heart Study.

**Figure 9.**
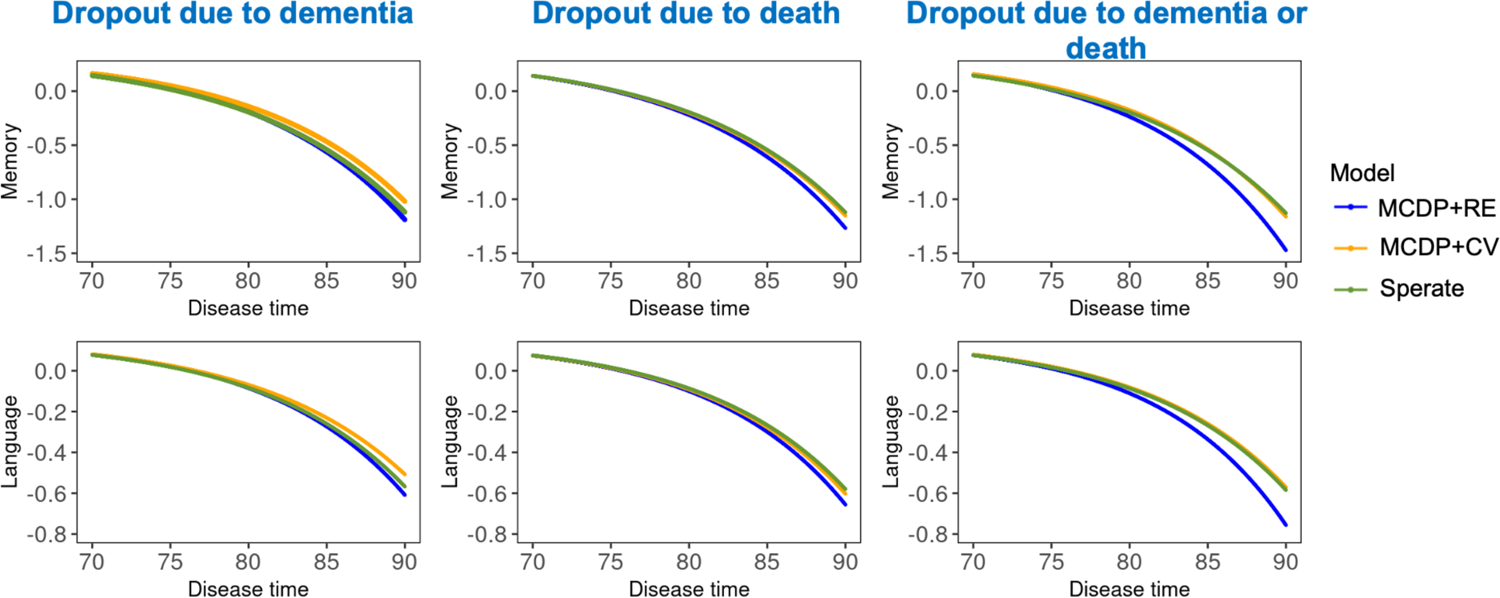
Mean disease progression trajectories fitted by joint models and separate models in the FHS. The left, middle, and right columns show the mean disease trajectories in scenarios where dropout is modeled as informative due to dementia, death, and both dementia and death, respectively. The y-axis in each panel represents the mean values of cognitive measure, and the x-axis denote the disease time estimated from the models. Three curves in each panel show trajectories fitted by different models: MCDP+RE (blue), MCDP+CV (orange), and separate models (green). MCDP+RE, the joint model with the shared random effect association structure; MCDP+CV, the joint model with the current value association structure; Separate, the separate models of the MCDP model and the proportional hazards model.

### Parameter interpretations

The parameter estimates of the four proposed joint models, including posterior means and 95% credible intervals (CI), as well as WAIC values were displayed in **Tables 8-10**. In all three scenarios, the WAIC values favored the MCDP+RE model, followed by the MCDP+CV model, indicating the real data likely exhibited a shared random effect data structure between the longitudinal and survival processes. Therefore, the following parameter interpretations were mainly based on the MCDP+RE model when both dementia and death were combined as a composite time-to-event outcome (**Table 10**). Longer education duration was associated with higher cognitive measures in both the memory and language domains (ॐ_)_ = 0.32, 95% CI: 0.28 to 0.37; ॐ_2_ = 0.15, 95% CI: 0.12 to 0.19). On average, women had a smaller disease age compared to men given the same actual chronological age (β = −2.11, 95% CI: −3.79 to −0.49). The standard deviation for the random latent time shift was estimated to be 8.45 years, implying substantial differences between actual chronological age and disease age for the FHS participants. Consequently, participants with similar actual chronological ages during follow-up may exhibit considerable variation in disease stages. The random intercepts showed a moderate positive relationship between the baseline values of the two cognitive domains, with an estimated correlation coefficient ρ_*b*_ of 0.57.

**Table 8.**
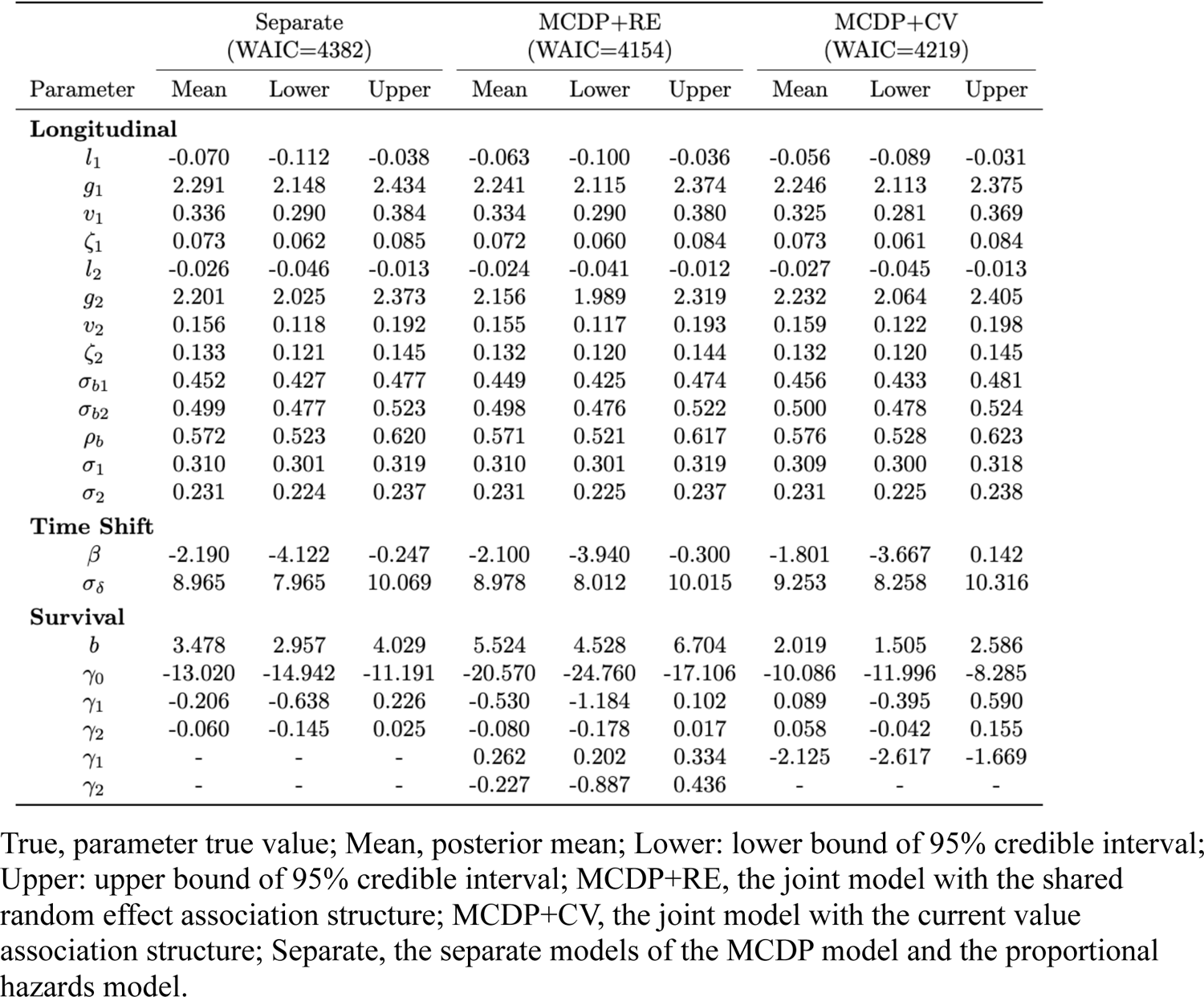
Real data application results when survival outcome is dementia onset.

**Table 9.** Real data application results when survival outcome is death.

| Parameter | Separate<br>(WAIC=5043) |  |  | MCDP+RE<br>(WAIC=5003) |  |  | MCDP+CV<br>(WAIC=5022) |  |  |
| --- | --- | --- | --- | --- | --- | --- | --- | --- | --- |
|  | Mean | Lower | Upper | Mean | Lower | Upper | Mean | Lower | Upper |
| **Longitudinal** |  |  |  |  |  |  |  |  |  |
| $l_1$ | -0.070 | -0.117 | -0.037 | -0.067 | -0.108 | -0.035 | -0.071 | -0.115 | -0.039 |
| $g_1$ | 2.293 | 2.152 | 2.446 | 2.245 | 2.097 | 2.397 | 2.288 | 2.148 | 2.436 |
| $v_1$ | 0.336 | 0.290 | 0.385 | 0.334 | 0.287 | 0.382 | 0.338 | 0.291 | 0.385 |
| $\zeta_1$ | 0.073 | 0.062 | 0.085 | 0.073 | 0.062 | 0.085 | 0.073 | 0.061 | 0.085 |
| $l_2$ | -0.026 | -0.046 | -0.012 | -0.025 | -0.043 | -0.012 | -0.026 | -0.046 | -0.012 |
| $g_2$ | 2.196 | 2.019 | 2.372 | 2.152 | 1.978 | 2.326 | 2.186 | 2.010 | 2.366 |
| $v_2$ | 0.155 | 0.117 | 0.193 | 0.153 | 0.114 | 0.192 | 0.154 | 0.117 | 0.193 |
| $\zeta_2$ | 0.133 | 0.121 | 0.145 | 0.133 | 0.121 | 0.145 | 0.133 | 0.121 | 0.145 |
| $\sigma_{b1}$ | 0.452 | 0.426 | 0.478 | 0.451 | 0.427 | 0.477 | 0.450 | 0.425 | 0.476 |
| $\sigma_{b2}$ | 0.498 | 0.476 | 0.522 | 0.498 | 0.476 | 0.521 | 0.498 | 0.475 | 0.521 |
| $\rho_b$ | 0.571 | 0.523 | 0.618 | 0.569 | 0.516 | 0.618 | 0.569 | 0.518 | 0.619 |
| $\sigma_1$ | 0.310 | 0.301 | 0.320 | 0.310 | 0.301 | 0.319 | 0.310 | 0.301 | 0.320 |
| $\sigma_2$ | 0.231 | 0.224 | 0.237 | 0.231 | 0.224 | 0.237 | 0.230 | 0.224 | 0.237 |
| **Time Shift** |  |  |  |  |  |  |  |  |  |
| $\beta$ | -2.193 | -4.167 | -0.303 | -2.198 | -4.081 | -0.368 | -2.246 | -4.246 | -0.340 |
| $\sigma_\delta$ | 8.980 | 7.936 | 10.126 | 8.668 | 7.682 | 9.716 | 8.945 | 7.952 | 10.028 |
| **Survival** |  |  |  |  |  |  |  |  |  |
| $b$ | 3.827 | 3.412 | 4.258 | 4.260 | 3.765 | 4.791 | 3.496 | 3.061 | 3.942 |
| $\gamma_0$ | -12.981 | -14.436 | -11.537 | -14.392 | -16.190 | -12.741 | -12.210 | -13.687 | -10.794 |
| $\gamma_1$ | -0.539 | -0.853 | -0.214 | -0.574 | -0.923 | -0.232 | -0.448 | -0.775 | -0.123 |
| $\gamma_2$ | -0.007 | -0.065 | 0.050 | -0.005 | -0.068 | 0.055 | 0.024 | -0.038 | 0.085 |
| $\alpha_1$ | - | - | - | 0.080 | 0.046 | 0.116 | -0.416 | -0.638 | -0.199 |
| $\alpha_2$ | - | - | - | 0.011 | -0.412 | 0.443 | - | - | - |

True, parameter true value; Mean, posterior mean; Lower: lower bound of 95% credible interval; Upper: upper bound of 95% credible interval; MCDP+RE, the joint model with the shared random effect association structure; MCDP+CV, the joint model with the current value association structure; Separate, the separate models of the MCDP model and the proportional hazards model.

**Table 10.** Real data application results when survival outcome is the composite of dementia and death.

| Parameter | Separate<br>(WAIC=5676) |  |  | MCDP+RE<br>(WAIC=5433) |  |  | MCDP+CV<br>(WAIC=5535) |  |  |
| --- | --- | --- | --- | --- | --- | --- | --- | --- | --- |
|  | Mean | Lower | Upper | Mean | Lower | Upper | Mean | Lower | Upper |
| **Longitudinal** |  |  |  |  |  |  |  |  |  |
| $l_1$ | -0.070 | -0.113 | -0.039 | -0.055 | -0.088 | -0.031 | -0.058 | -0.095 | -0.031 |
| $g_1$ | 2.290 | 2.148 | 2.440 | 2.151 | 2.023 | 2.288 | 2.222 | 2.090 | 2.365 |
| $v_1$ | 0.336 | 0.290 | 0.385 | 0.324 | 0.283 | 0.368 | 0.328 | 0.285 | 0.373 |
| $\zeta_1$ | 0.073 | 0.061 | 0.085 | 0.072 | 0.061 | 0.084 | 0.072 | 0.061 | 0.084 |
| $l_2$ | -0.026 | -0.045 | -0.012 | -0.022 | -0.038 | -0.010 | -0.026 | -0.044 | -0.013 |
| $g_2$ | 2.189 | 2.003 | 2.365 | 2.085 | 1.907 | 2.256 | 2.193 | 2.031 | 2.369 |
| $v_2$ | 0.154 | 0.116 | 0.193 | 0.151 | 0.115 | 0.189 | 0.157 | 0.118 | 0.196 |
| $\zeta_2$ | 0.133 | 0.121 | 0.144 | 0.132 | 0.121 | 0.145 | 0.132 | 0.120 | 0.145 |
| $\sigma_{b1}$ | 0.452 | 0.428 | 0.477 | 0.450 | 0.427 | 0.473 | 0.453 | 0.429 | 0.478 |
| $\sigma_{b2}$ | 0.499 | 0.478 | 0.523 | 0.497 | 0.476 | 0.520 | 0.499 | 0.477 | 0.522 |
| $\rho_b$ | 0.572 | 0.522 | 0.619 | 0.569 | 0.518 | 0.615 | 0.572 | 0.519 | 0.621 |
| $\sigma_1$ | 0.310 | 0.302 | 0.320 | 0.308 | 0.299 | 0.317 | 0.308 | 0.299 | 0.318 |
| $\sigma_1$ | 0.230 | 0.224 | 0.237 | 0.231 | 0.225 | 0.237 | 0.231 | 0.225 | 0.238 |
| **Time Shift** |  |  |  |  |  |  |  |  |  |
| $\beta$ | -2.214 | -4.134 | -0.307 | -2.115 | -3.794 | -0.485 | -1.966 | -3.811 | -0.066 |
| $\sigma_\delta$ | 8.958 | 7.946 | 10.111 | 8.451 | 7.600 | 9.389 | 9.002 | 8.036 | 10.065 |
| **Survival** |  |  |  |  |  |  |  |  |  |
| $b$ | 3.717 | 3.382 | 4.053 | 4.846 | 4.311 | 5.415 | 2.970 | 2.612 | 3.339 |
| $\gamma_0$ | -12.370 | -13.501 | -11.247 | -16.105 | -18.056 | -14.305 | -10.730 | -11.943 | -9.573 |
| $\gamma_1$ | -0.425 | -0.674 | -0.170 | -0.543 | -0.885 | -0.205 | -0.224 | -0.487 | 0.044 |
| $\gamma_2$ | -0.024 | -0.072 | 0.025 | -0.025 | -0.078 | 0.028 | 0.048 | -0.005 | 0.098 |
| $\alpha_1$ | - | - | - | 0.154 | 0.120 | 0.190 | -1.024 | -1.263 | -0.783 |
| $\alpha_2$ | - | - | - | -0.070 | -0.429 | 0.307 | - | - | - |

True, parameter true value; Mean, posterior mean; Lower: lower bound of 95% credible interval; Upper: upper bound of 95% credible interval; MCDP+RE, the joint model with the shared random effect association structure; MCDP+CV, the joint model with the current value association structure; Separate, the separate models of the MCDP model and the proportional hazards model.

In the survival sub-model, women had a smaller hazard of developing dementia or death than men after the adjustment for other factors (HR=0.58, 95% CI: 0.41 to 0.81). Longer education duration lowered the risk of dementia or death, although this association was not significant (HR=0.98, 95% CI: 0.92 to 1.03). In addition, the significant positive association between the random latent time shift and the hazard of dementia or death confirmed that dropouts due to dementia or death represented informative missingness. Specifically, for two participants with the same actual chronological age, sex, and education level, the one with a greater disease age faced a higher hazard of informative dropout (HR=1.17, 95% CI: 1.13 to 1.21). There was no evidence of a significant association between the random intercept and the hazard of dementia or death (HR=0.93, 95% CI: 0.65 to 1.36). When the data was modeled by the MCDP+CV model, the significant negative association between the current value of cognitive measures and the hazard of dropout also indicated informative missingness in the longitudinal measures, consistent with the findings in the MCDP+RE model.

### Model comparison

The WAIC values suggested that the models that account for informative missingness outperformed the separate models, with the MCDP+RE model showing the best predictive accuracy in all three scenarios. The MCDP+RE model showed the most pronounced decreasing trend for both cognitive domains in all three scenarios (Figure 9), while the other two models produced more optimistic estimates with slower rates of decline in their trajectories. Note that when either dementia or death was considered as the only survival outcome, three models demonstrated similar mean trajectories for disease progression. Incorporating both dementia and death as outcomes caused the mean disease progression trajectories in the MCDP+RE model to diverge more substantially from the other two models, due to an increase in the event rate. These findings were consistent with the simulation results.

## 6. Discussion

In this project, we developed a joint model that integrates a multivariate nonlinear disease progression model with latent time shifts. To evaluate the model’s capacity to handle informative dropout in longitudinal data, we compared joint models with two association structures and the separate models that ignore informative missingness under a range of simulation scenarios.

Specifically, we considered the joint models that account for informative dropout by modeling the associations between the longitudinal and dropout processes, i.e., the shared random effect structure and the current value structure. We also conducted the separate models that ignored informative dropout, where the longitudinal and dropout processes were modeled independently. Our simulation showed that all three models incorporating latent time shifts can realign individual trajectories along a new global disease timeline. In addition, our proposed joint models can effectively account for informative dropout in the longitudinal data compared to the disease progression model in the separate models. Specifically, in the presence of informative dropout, the joint models with correctly specified association structures outperformed the others, providing the lowest bias and the most precise estimates of mean disease progression trajectories and latent time shifts. Even the joint model with a misspecified association structure captured the relationship between two processes and yielded intermediate results, offering advantages over separate models. Compared to the joint models, the separate models tended to provide over-optimistic estimates with respect to mean disease progression. Although varying the sample size, the span of latent time shifts, or the event rate could influence model performance, the improvement of joint models over separate models consistently remained robust. When dropout was unrelated to the longitudinal data, i.e., non-informative, all joint and separate models performed similarly, showing unbiased results.

The application using NP test scores and time to dementia or death in the FHS Offspring cohort confirmed our simulation findings regarding model comparison. WAIC values suggested that the relationship between longitudinal cognitive measures and time to dementia or death may resemble a shared random effect association structure. The significant non-zero association parameters in the joint models confirmed that dropouts due to dementia or death were informative, aligning with the findings from WAIC values. Furthermore, we showed sex influenced the latent time shift and the risk of dementia or death, while education was associated with the average cognitive levels.

Our findings were consistent with prior studies. First, most of the previous studies on joint models, both linear and nonlinear, showed that joint models outperformed separate models in the presence of informative dropout [24–28, 30, 32], which was consistent with our key findings. Second, the mix-effects model in the separate models that ignored informative dropout showed over-optimistic estimates for the disease progression trend, corresponding to conclusions drawn in previous studies [26–28]. It was shown that subjects who dropped out due to excessively high or low longitudinal measures were downweighed during the estimation procedure, so they contributed weakly to the likelihood function in the mixed-effects models. For example, mixed-effects models for cognitive decline might overestimate the rate of disease progression over time. This overestimation occurs because participants with lower cognitive scores, who are more likely to drop out, have a reduced influence on the model’s estimation process due to informative missingness. Therefore, failing to simultaneously consider the dropout process can lead to biased. Third, applications of joint models in the dementia studies revealed that the hazard of dementia or death was associated with the true unobserved values of cognitive measures [29, 30]. This finding corresponded to the results in our joint models with the current value structure. Our study has some limitations. First, the MCDP model assumes an exponential curve for longitudinal cognitive measures, which represents a monotonic decreasing trend over time.

However, this assumption may not apply to short-term observations, as participants can show cognitive improvements within months [48]. Patients can revert from mild cognitive impairment (MCI) to normal cognition and then transition back to MCI in the short term. Consequently, the assumption of a monotonic exponential curve fits our data, which were collected every four to five years, but may not generalize well to short-term scenarios. Second, we considered time to dementia or death as a composite survival outcome instead of modeling them as competing or semi-competing risks. Previous studies showed the competing risks joint model that distinguishes between two causes of dropout may outperform the standard joint model that treats all the dropouts due to different causes equally in general [26, 30, 32]. However, this improvement is most pronounced only when two types of dropouts have substantially different relationships with the longitudinal measures, e.g., one type of dropout is informative and the other one is non-informative, or two competing risks are both informative but oppositely associated with the longitudinal measures. In our application, both dropouts are informative and exhibit the same directionality for their associations with the longitudinal process. Thus, ignoring competing risks in the joint model estimation has minimal influence on our conclusions.

Our study has come strengths. First, our proposed joint model is an extension of a previously proposed multivariate nonlinear mixed-effects model with latent time shifts for disease progression [3]. To the best of our knowledge, this is the first integration of such a disease progression model into a joint model framework. Previous disease progression models with latent time shifts treat missing data as MAR, leading to potential biased results. Our proposed joint model corrects the bias by using the latent time shifts as a tool to simultaneously model the dropout process. Second, we compared different association structures within the joint model framework with the separate models of longitudinal and dropout processes across various data structure scenarios. Few prior studies compared the performance of multivariate nonlinear joint models using both shared random effect and current value association structures. Third, previous disease progression models with latent time shifts were primarily applied to specific disease cohorts, like the Alzheimer’s Disease Neuroimaging Initiative (ADNI) [3, 9, 49]. In contrast, we evaluated our model using a community cohort, offering new insights into dementia progression, including Alzheimer’s disease, within a general population.

Our study can be extended in the following ways. First, our proposed joint model can be further adapted to account for different shapes of dementia progression curves, such as sigmoid or quadratic functions, for conducting sensitivity analyses [10]. Other models have postulated a changepoint during disease progression as an alternative to describe the accelerated rate of cognitive decline at a later disease stage [50–52]. Second, our model can be extended to distinguish dropouts due to dementia, death, and other causes by treating them as competing risks or semi-competing risks. Joint models with distinct reasons for dropout not only correct bias in the longitudinal process but also assess the impact of longitudinal process on each dropout type [26, 30, 32]. Third, in aging research, dementia onset is usually treated as interval-censored [12, 53]. Our joint model can be extended to account for interval censoring, which may help reduce bias in estimating both longitudinal and dropout processes. For example, Gueorguieva et al. (2012) considered interval-censored dropout times in a joint model for competing risks and distinguished various dropout causes [26].

In conclusion, we incorporated a multivariate nonlinear mixed-effects model for disease progression with latent time shifts into a joint model framework. To handle informative missingness in the longitudinal data, we evaluated and compared different association structures within the joint models. In addition, we compared the joint models with the separate models that ignored informative dropout. We demonstrated the improvement of joint models over separate models across various scenarios. Future extensions could explore alternative parametrizations of the longitudinal sub-model and incorporate competing risks with interval censoring into the survival sub-model.

## Data Availability

All data produced in the present study are available upon reasonable request to the authors

